# Metagenomic Analysis Reveals A Possible Association Between Respiratory Infection and Periodontitis

**DOI:** 10.1101/2020.12.08.20244723

**Authors:** Zhenwei Liu, Tao Zhang, Keke Wu, Zhongshan Li, Xiaomin Chen, Shan Jiang, Lifeng Du, Saisai Lu, Chongxiang Lin, Jinyu Wu, Xiaobing Wang

## Abstract

Periodontitis is an inflammatory disease which is characterized by progressive destruction of the periodontium and causes tooth loss in adults. Periodontitis is known to be associated with dysbiosis of the oral microflora, often linked to various diseases. However, the complexity of plaque microbial communities of periodontitis, and antibiotic resistance and enhanced virulence make this disease difficult to treat. Therefore, we used metagenomic shotgun sequencing in this study to investigate the etiology, antibiotic-resistant genes (ARGs) and virulence genes (VirGs) of periodontitis. We revealed a significant shift in the composition of oral microbiota as well as several functional pathways that were represented significantly more abundant in periodontitis patients than in controls. Additionally, we observed several positively selected ARGs and VirGs with the Ka/Ks ratio > 1 by analyzing our data and a previous periodontitis dataset, indicating that ARGs and VirGs in oral microbiota may suffer from positive selection. Moreover, 5 of 12 positively selected ARGs and VirGs in periodontitis patients were found in the genomes of respiratory tract pathogens. Of note, 91.8% of the background VirGs with at least one non-synonymous single-nucleotide polymorphism for natural selection were also from respiratory tract pathogens. These observations suggest a potential association between periodontitis and respiratory infection at the gene level. Our study enriches the knowledge of pathogens and functional pathways as well as the positive selection of antibiotic resistance and pathogen virulence in periodontitis patients, and provides evidence from the gene level for an association between periodontitis and respiratory infection.

## Introduction

Periodontitis is an inflammatory disease of the periodontium which is characterized by progressive destruction of the periodontium, and causes tooth loss in adults. It is one of the most well-characterized human diseases associated with dysbiosis of the oral microflora [1]. The inflammatory infiltration induced by the microbes often destroys the supportive tissue, leading to alveolar bone loss before developing into an irreversible situation [2, 3]. A number of systemic diseases have been reported to be associated with periodontitis, such as atherosclerotic vascular disease, cardiovascular disease, adverse pregnancy outcomes, Alzheimer’s disease, rheumatoid arthritis, colorectal cancer, and diabetes [4-10]. Moreover, periodontitis pathogens of the oral cavity as well as respiratory pathogens have been found to may access the respiratory tract and induce respiratory infection [11, 12].

Among the factors contributing to the periodontitis, virulence factors of the oral microbiota have been a critical and important issue, influencing the onset of the disease. A set of key virulence factors produced by of *Porphyromonas gingivalis*, a well-established oral keystone pathogen, had been discovered and determined for the role of pathogenesis, such as Lys-and Arg-proteases, lipoprotein and atypical lipopolysaccharide [13]. Bacterial sialidases have been considered virulence factors in oral pathogens like *Tannerella forsythia* [14, 15]. In addition, functional studies showed the activation of the metalloproteases, peptidases, and proteins involved in iron metabolism in oral pathogens might be linked to the potential association with periodontal disease [16, 17].

Antibiotic resistance, described as “bacteria changing in ways that reduce or eliminate the effectiveness of antibiotics”, is another impediment to treatment [18]. When drug-sensitive pathogens are eliminated, drug-resistant pathogens are left to be selected [19]. Bacteria often acquire resistance by random mutations in functional genes or by horizontal gene transfer (HGT). The exchange of genes via HGT is common among bacteria and can facilitate the spread of drug resistance [18]. Considering that the evolutionary dynamics of drug resistance is usually derived from the increasing overuse of antibiotics [20], the empirical and broad use of antibiotics resulting from the lack of clarification of periodontal disease pathogens may increase the risk of antibiotic-resistance problems in periodontitis [21]. An increasing number of studies have reported that subgingival periodontal pathogens from chronic periodontitis patients are resistant to at least one of the commonly used antibiotics, and many strains from the “red complex” have already developed resistance to various antibiotics [22, 23]. Therefore, it is necessary to gain more insight into drug-resistance genes and associated pathways in periodontitis.

In this study, we explored the difference in composition and functional pathways of oral microbiota between patients with periodontitis and individuals with a healthy periodontium using metagenomic shotgun sequencing. We also investigated potential bacteria, virulence genes (VirGs) and antibiotic resistance genes (ARGs) associated with periodontitis. Our results contribute to enriching our understanding of the bacterial ecology of periodontitis and reveal a potential association between periodontitis and respiratory infections.

## Results

### Metagenomic sequencing and data quality control

A total of 30 patients with periodontitis and 15 healthy individuals were recruited with clinical information including age, sex, diagnosis and tooth numbers (Table S1). Due to the insufficient amount of DNA extracted from each dental plaque sample, every five samples were pooled together, and six periodontitis specimens (P1, P2, P3, P4, P5 and P6) and three controls (C1, C2 and C3) were ultimately obtained. Sequencing of the six periodontitis specimens and three controls generated 2.38–2.68 gigabases (Gb) of raw sequencing data consisting of 30–32 million pair end (PE) reads per sample (Table S2). After data processing, 2.30–2.50 Gb of clean high-quality data was retained, consisting of 29–31 million PE reads for each sample. The clean read rate for each sample ranged from 95% to 97% (Figure S1A), and the clean Q30 base rate ranged from 92% to 93%. The low-quality read rate, NS read (read with “N” or “.” bases that couldn’t be assign a specific base by the basecaller) rate, and adapter read rate were as low as expected (Figure S1B and Table S2). Alignment against hg19 revealed that the human sequence read rate ranged from 74% to 88% for all specimens (Figure S1A). After removal of human sequence contamination, we retained approximately 3.58–7.89 million reads for all samples for subsequent analyses (Table S2).

### Significant differences in the composition and diversity of oral microbiota between periodontal patients and controls

To explore the taxonomic composition, we applied MetaPhlAn2 [24] to analyze the abundance information of each sample in each taxonomy hierarchy. At the kingdom level, the analysis revealed almost all the reads aligned to Bacteria, 0.16% reads aligned to viruses, and 0.06% reads aligned to Archaea (Table S3). At the phylum level, there were ten phyla aligned, among which *Bacteroidetes* was the most dominant bacterial phylum (**Figure 1**A). Both *Synergistetes* (Wilcoxon rank sum test, *P* = 0.024) and *Spirochaetes* (Wilcoxon rank sum test, *P* = 0.024) significantly dominated in periodontal patients over controls (Figure 1B and Table S4). At the genus level, 69 genera were aligned, and the top 30 genera aligned 89–97% of reads in all samples. Among these genera, *Prevotella* was the most dominant genus in periodontal patients. There were *Treponema, Desulfobulbus, Fretibacterium, Tannerella*, and *Anaeroglobus* dominant in periodontal patients, and *Corynebacterium* dominant in controls (Wilcoxon rank sum test, *P* < 0.05) (Figure S2A and Table S5). At the species level, rarefaction curves indicated that sequence data could be sufficient to identify the majority of species (Figure S2B) and the read alignment identified a total of 168 species, among which there were five species enriched in periodontal patients, three species dominant in controls (Wilcoxon rank sum test, *P* < 0.05) (Figure 1C and Table S6).

**Figure 1.**
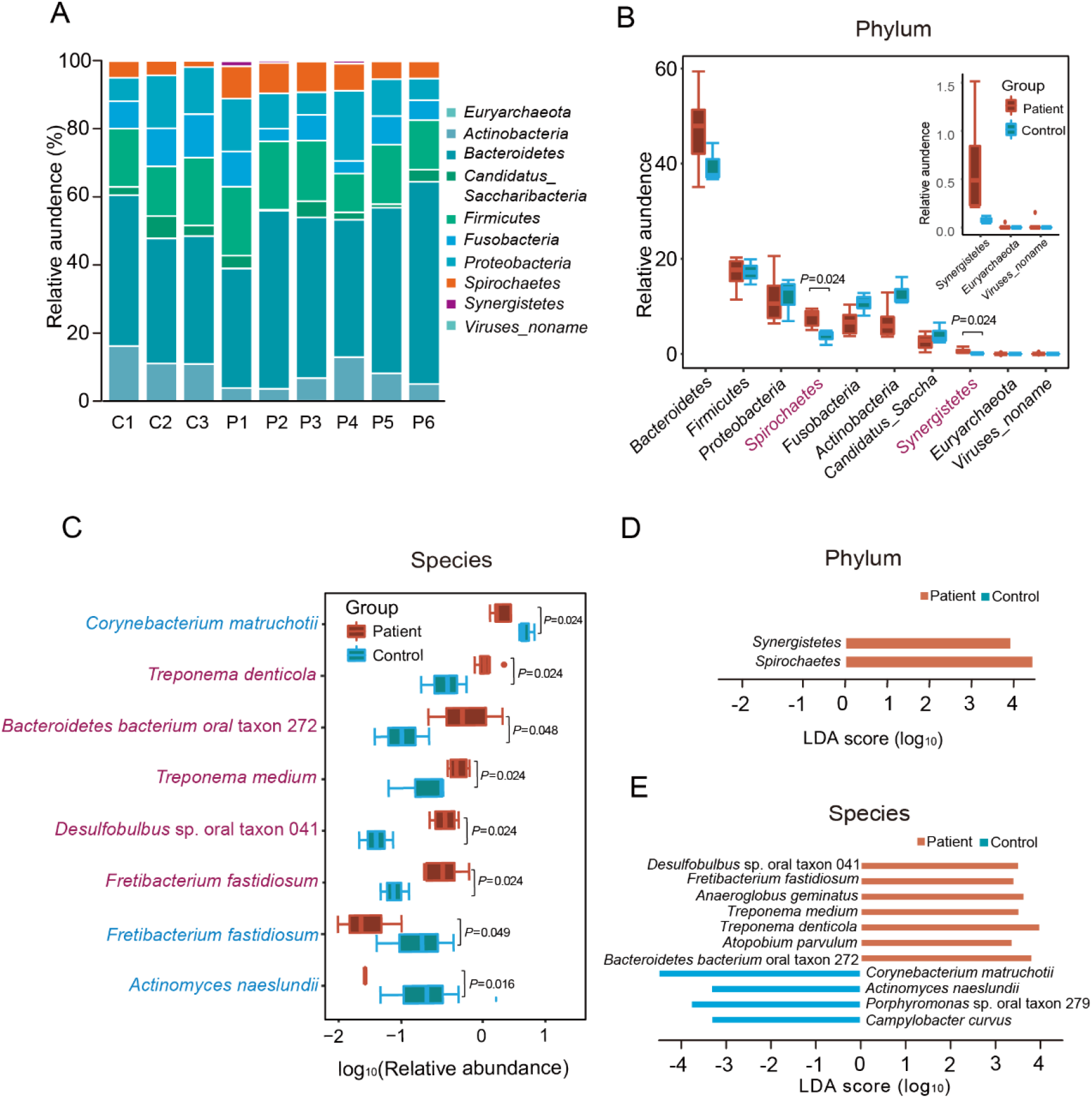
The shift in oral microbiota composition at the phylum and species level. **A**. Composition of oral microbiota at the phylum level in each sample. **B**. Comparison of each phylum between periodontitis and control samples. The significantly abundant phyla in periodontitis samples are marked with red. The three small boxplots in the boxplot-in-boxplot indicate a zooming state for its corresponding boxplot in a vertical direction in the main boxplot. **C**. Eight species with significant difference between periodontitis and control samples. The significantly abundant species are marked with red (in periodontitis samples) or light blue (in control samples). **D**. LEfSe analysis results at the phylum level. **E**. LEfSe analysis at the species level.

LEfSe analysis at the phylum level revealed linear discriminant analysis (LDA) scores of *Synergistetes* and *Spirochaetes* greater than 3 in periodontal patients (Figure 1D). At the genus level, LEfSe analysis showed that *Treponema, Desulfobulbus*, and *Fretibacterium* in periodontal patients and *Corynebacterium* in controls had an LDA score passing the threshold of 3 (Figure S2C). At the species level, *Treponema denticola, Treponema medium, Fretibacterium fastidiosum, Desulfobulbus* sp. oral taxon 041, *Bacteroidetes bacterium* oral taxon 272, *Anaeroglobus geminatus* and *Atopobium parvulum* in periodontitis patients, as well as *Corynebacterium matruchotii, Actinomyces naeslundii, Porphyromonas* sp. oral taxon 279 and *Campylobacter curvus* in controls, passed the LDA cut-off threshold of 3 (Figure 1E).

For beta diversity, principal component analysis (PCA) and non-metric multidimensional scaling (NMDS) analysis were performed at the phylum and species levels (Figure S3A and B; **Figure 2**A and B). Both PCA and NMDS showed distinct distribution of distance between periodontitis patients and controls, although there was no significant difference. Similar observation was shown up in the analysis of similarities (ANOSIM) of the NMDS analysis at the phylum and species levels (Figure S3C and Figure 2C). For alpha diversity at the phylum level, the Shannon index, Simpson index, InverseSimpson index, and Pielou index tended to be higher in controls than in periodontitis patients but without statistical significance (Figure S3D–G). At the species level, the Shannon index and Pielou index were significantly higher in periodontitis patients than in controls (Figure 2D and E). The Simpson index and InverseSimpson index at the species level also tended to be higher in periodontitis patients, although not significantly (Figure S3H and I).

**Figure 2.**
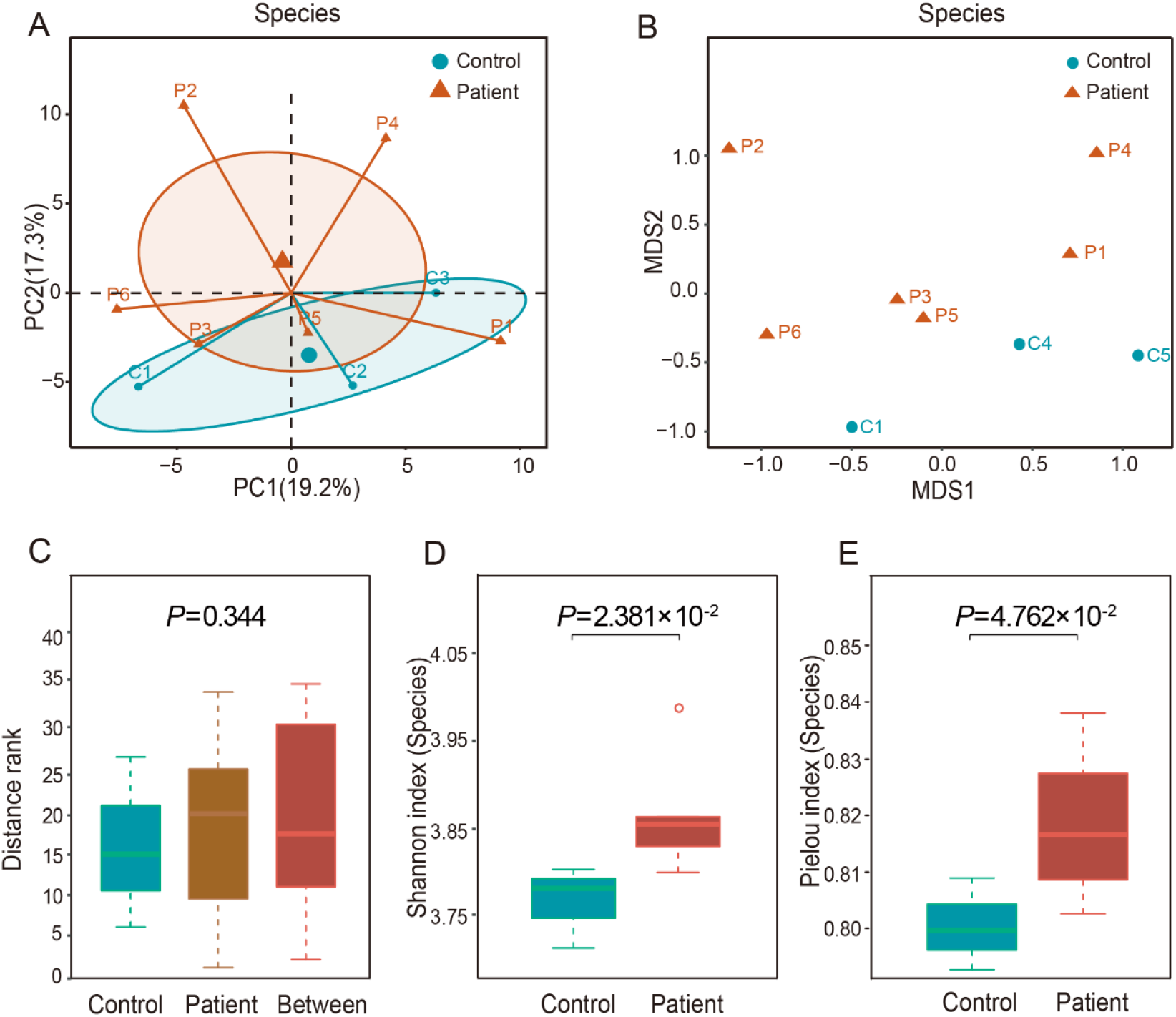
Alpha diversity and beta diversity at the species level. **A**. Principal component analysis (PCA) at the species level. **B**. the non-metric multidimensional scaling (NMDS) analysis of all samples at the species level. **C**. the analysis of similarities (ANOSIM) of NMDS results at the species level. Both the Shannon index (**D**) and Pielou index (**E**) of the alpha diversity at the species level. The two indexes were significantly higher in periodontitis samples.

### Functional annotation associated with drug resistance and virulence is significantly enriched in periodontitis

To determine the differential function of microbial taxa between periodontitis patients and controls, we performed the several functional predictions including eggNOG annotations, Gene Ontology (GO) and Kyoto Encyclopedia of Genes and Genomes (KEGG) pathway analysis, as well as the corresponding gene set enrichment analysis (GSEA). According to the annotations with a false discovery rate (FDR) < 0.05 after applying DESeq2 with reads per kb (RPK) values, we found 22 EggNOG terms significantly abundant in periodontitis patients, among which there were ten terms associated with transposase that might function as resistance against many different antibiotics. Several EggNOG terms functioning in strengthening physical defenses were also up-regulated in periodontitis, including the multidrug and toxin compound extrusion (MATE) efflux family protein, the ATP-binding cassette (ABC) superfamily transporter extruding multidrug and toxic compounds, and cell wall/membrane/envelope biogenesis [25] (**Figure 3**A and Table S7). GSEA of EggNOG terms revealed several significantly enriched terms (FDR < 0.05, normalized enrichment score [NES] > 1) in periodontitis patients. Several transposase terms and ABC-transporters terms were still significantly enriched in periodontitis patients indicating their important role in periodontitis. Several terms involved in the synthesis of fatty acid were found significantly enriched in controls, such as fatty acid synthase, esterase and MaoC domain protein dehydratase as well as oxidoreductase (**Figure 3**B and Table S8).

**Figure 3.**
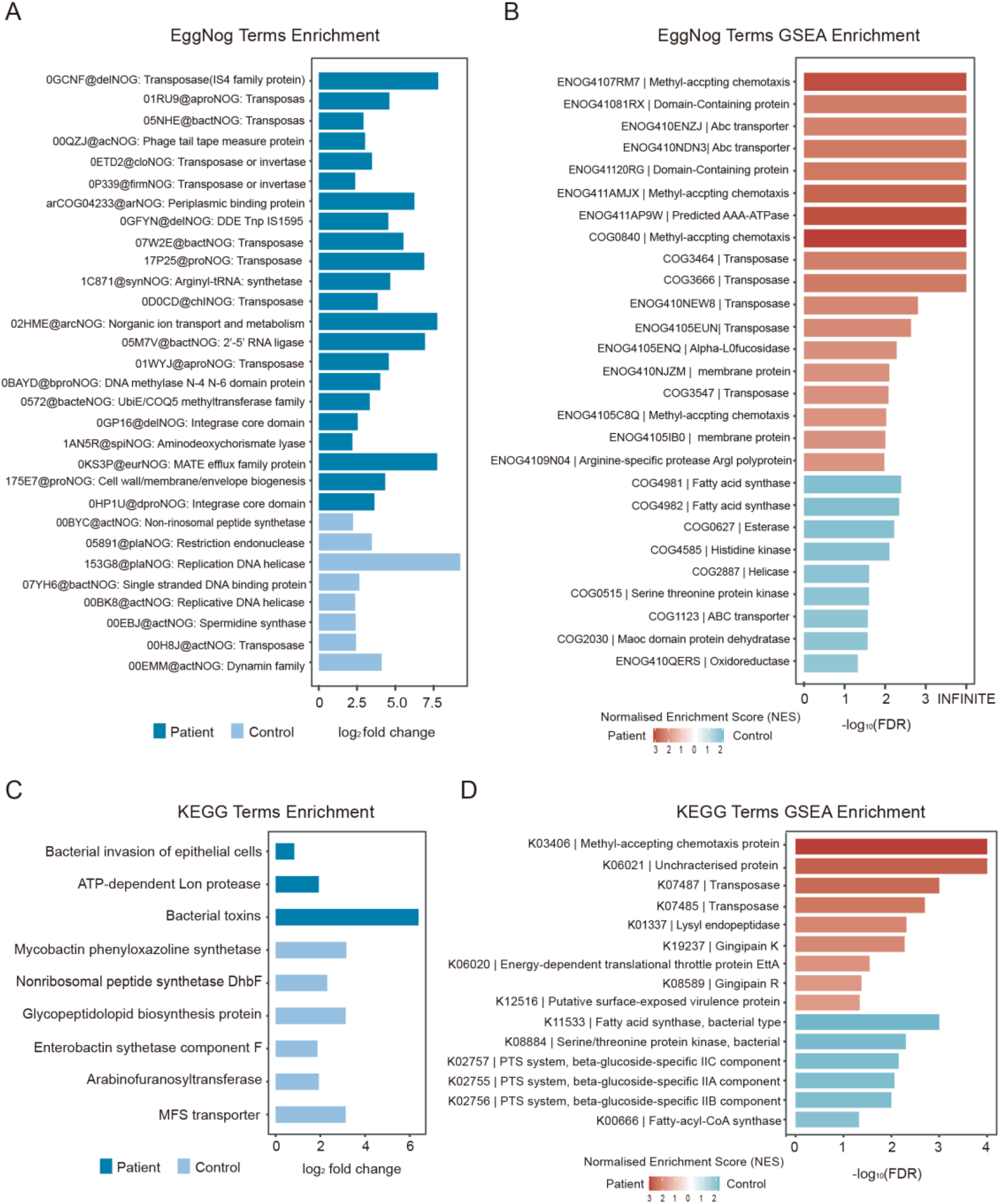
Significant differences between periodontitis and control samples in various functional annotations. **A**. Significantly abundant EggNOG terms in periodontitis samples or control samples with false discovery rate (FDR) < 0.05 and log_2_ fold change (log_2_(FC)) > 0.7. **B**. Gene set enrichment analysis (GSEA) results of EggNOG terms with normalized enrichment score (NES) > 1 and FDR < 0.05 in periodontitis patients and controls. **C**. Significantly abundant KEGG terms in periodontitis samples or control samples (FDR < 0.1, log_2_(FC) > 0.7). **D**. GSEA results of KEGG terms with NES > 1 and FDR < 0.05 in periodontitis patients and controls

The analysis of GO terms showed that there were 39 GO terms were down-regulated in periodontitis patients (Table S7). GSEA of GO terms revealed no significantly enriched terms in periodontitis patients but found 75 GO terms significantly enriched in controls, including synthesis of fatty acids, cell cycle processes and membranes as well as several other GO terms which may all contribute to the maintenance of health status (Table S8). There were three KEGG pathways with an FDR < 0.1 increased in periodontitis patients, including bacterial invasion of epithelial cells, ATP-dependent long proteases, and bacterial toxins (Figure 3C and Table S7). GSEA of KEGG terms revealed several significantly enriched proteins in periodontitis patients identical to GSEA of EggNOG terms, such as methyl-accepting chemotaxis protein (MCP), transposase as well as unique virulence proteins. In controls, one KEGG term (fatty acid synthase, bacteria type) with significant enrichment was consistent with the GSEA enrichment for EggNOG terms (Figure 3D and Table S8).

### ARG and virulence annotation revealed convergent enrichment in several genes in periodontitis from a significantly different gene context

To investigate the abundances of each gene family in the oral microbiota from periodontitis patients and controls, we use the HUMAnN2 to annotate all sequencing reads and found 196 gene families significantly abundant in periodontitis patients. Several genes encode various functional type of protein, including resistance protein, transposon, transposase or transporters (Table S7). For example, A3CPG0 was annotated as an ABC-type bacitracin resistance protein with a taxon assigned to *Streptococcus sanguinis*. F0H8B2 and F2KUK8 were the conjugative transposon proteins (TraA) encoded in *Prevotella denticola*. F5RK90 was a transposase from *Centipeda periodontii*. S8FC11 and S8GNT2 were ABC transporters that function in *Bacteroidetes bacterium* oral taxon 272. C8PRE7 and C9LSP5 were two flagellar FlbD family proteins encoded in *Treponema vincentii* ATCC 35580 and *Selenomonas sputigena* that function in the invasion of pathogens. Most of the other genes were associated with translation, such as 30S and 50S ribosomal proteins and transcriptional regulators (Table S7).

To further explore the potential ARGs from the oral microbiota, we used DeepARG tool to annotate the reads [26], and found that there were 30 ARG biomarkers with LDA scores > 2 in periodontitis patients using LEfSe analysis (Table S9). For VirGs, after sequence alignment and annotation, LEfSe analysis revealed 10 VirG biomarkers with LDA scores > 2 in periodontitis (Table S9).

To eliminate sample bias, we analyzed the metagenomic data for dental plaque of periodontal health (PHP) and disease (PDP) from a previous study by Wang et al. as a comparison [27]. After removal of human DNA contamination, we found that there was no significant difference of non-human sequence rate either in periodontitis patient samples (Wilcoxon rank sum test, *P* = 5.594×10^−2^) or controls (Wilcoxon rank sum test, *P* = 0.161) between current dataset and study by Wang et al. (Table S10) [27]. The PCA and ANOSIM analysis using the relative abundance of genes aligned by HUMAnN2 showed that a significant difference between the two studies. This difference might be due to several factors including the difference in the regions where participants resided or technical bias for library construction and sequencing between two datasets, even though the same analytical methods were used (Figure S4A and B, *P* = 3.792×10^−2^). After ARG annotation, LEfSe analysis revealed 67 ARG biomarkers with LDA scores > 2 in periodontitis from the previous study (Table S11). For VirGs, LEfSe analysis revealed 70 VirG biomarkers with LDA scores > 2 in periodontitis from the previous study (Table S11). In addition, comparative analysis indicated that there were two identical ARGs as well as two identical VirGs between our dataset and dataset from the previous study (**Figure S4C and D**).

### MAF of non-synonymous SNPs in ARGs and VirGs are significantly higher in periodontitis

After SNP calling in ARGs from each sample by BCFtools and VarScan, we calculated the minor allele frequency (MAF) of non-synonymous SNPs and unique non-synonymous SNPs, as well as synonymous SNPs in ARGs and VirGs between periodontitis patients and controls from current dataset and study by Wang et al. [27]. The rarefaction curve of SNPs in ARGs and VirGs showed that SNP number was sufficient for analysis of all samples (Figure S5). We found that the MAF of non-synonymous SNPs in ARGs was significantly higher in periodontitis patients than controls in current study. The same difference was also observed in the previous study (**Figure 4**A-C). For unique non-synonymous ARG SNPs, similar results were also obtained in both studies (Figure 4D-F). However, there was no significant difference for the MAF of synonymous SNPs in ARGs between periodontitis patients and controls in either our or previous study (Figure S6A-C). These findings indicated that ARGs in periodontitis patients may suffer more selection pressure compared with those in controls. Similarly, we also calculated the MAFs of non-synonymous and unique non-synonymous SNPs in VirGs in periodontitis patients and controls from both studies. There was a significant difference in the MAF of non-synonymous (Figure 4G-I) and unique non-synonymous SNPs (Figure 4J-L) between periodontitis patients and controls, while there was no significant difference in the MAF of synonymous SNPs between periodontitis patients and controls in either study (Figure S6D–F). The MAF of SNPs in VirGs could indicate that VirGs may also be subject to positive selection in periodontitis patients compared with controls.

**Figure 4.**
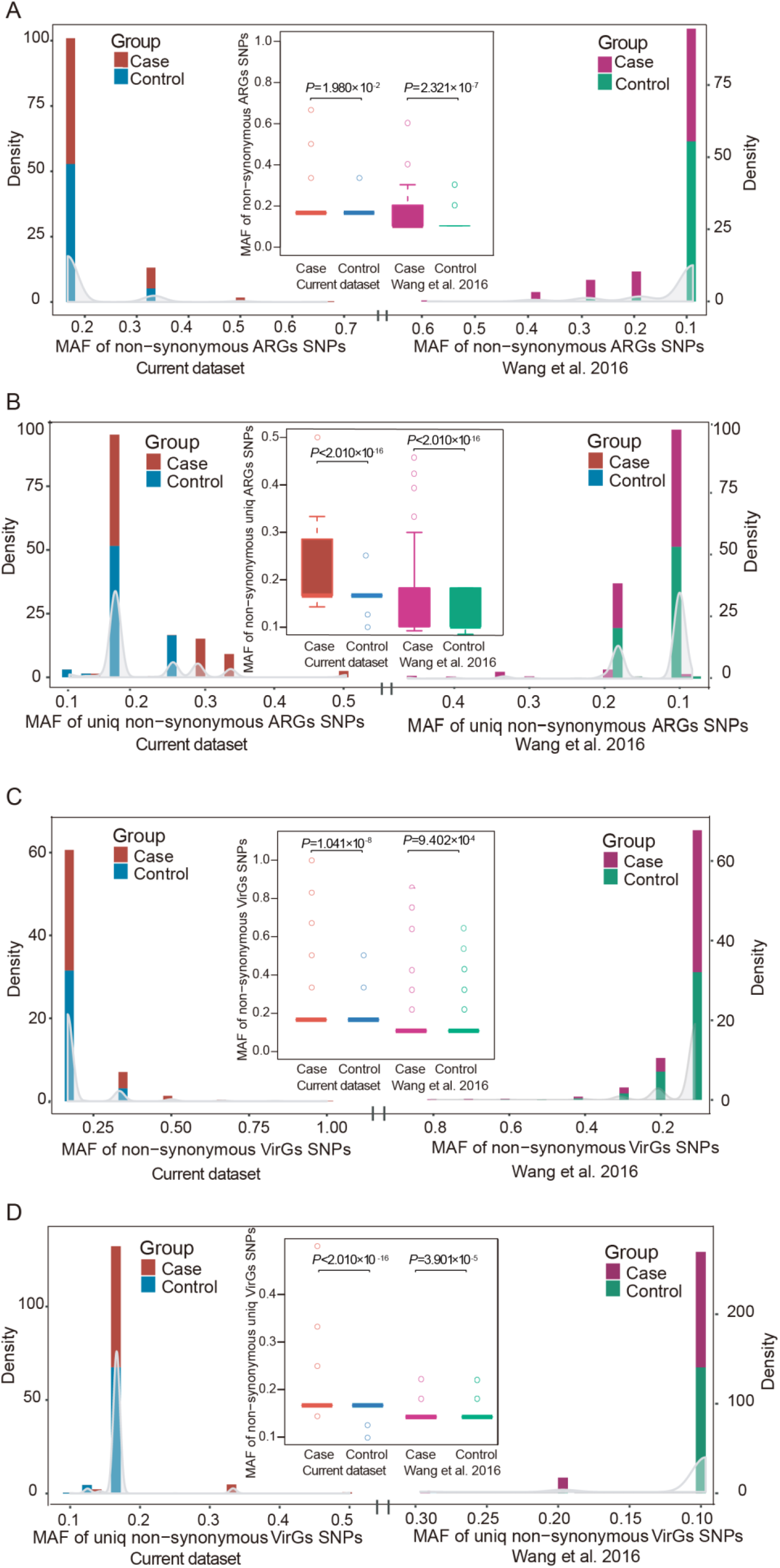
Comparison of non-synonymous SNPs in ARGs and VirGs between periodontitis and control samples from both datasets. **A**. Distribution of the minor allele frequency (MAF) of non-synonymous SNPs in ARGs from both current studyand previous study. Comparison of the MAF of non-synonymous SNPs in ARGs between periodontitis and control in both datasets. **B**. Distribution of the MAF of unique non-synonymous SNPs in ARGs from both current study and previous study. Comparison of the MAF of unique non-synonymous SNPs in ARGs between periodontitis and control in both datasets. **C**. Distribution of the MAF of non-synonymous SNPs in VirGs from both current study and previous study. Comparison of the MAF of non-synonymous SNPs in VirGs between periodontitis and control in both datasets. **D**. Distribution of the MAF of unique non-synonymous SNPs in VirGs from both current study and previous study. Comparison of the MAF of unique non-synonymous SNPs in VirGs between periodontitis and control in both datasets. All *P* values were calculated using Wilcoxon rank sum test. SNPs, single-nucleotide polymorphisms; ARGs, antibiotic resistance genes; VirGs, virulence genes.

### Several ARGs and all VirGs with Ka/Ks > 1 in periodontitis were frequent from respiratory infection pathogens

To identify positively selected ARGs, the Ka/Ks rate was calculated for each ARG. According to previous studies, a Ka/Ks ratio > 1 indicates positive selection and conversely, a Ka/Ks ratio < 1 implies negative selection [28, 29]. In our dataset, there were six ARGs with a Ka/Ks rate > 1 in periodontitis and a Ka/Ks rate < 1 in controls including *macB* (*Yersinia enterocolitica, Faecalibacterium* sp. CAG:82, and *beta proteobacterium* AAP51), *mexD* and *cpxR* (*Pseudomonas aeruginosa*), and *pgpB* (*Porphyromonas gingivalis*) (Table S12 and **Figure 5**A). These genes were considered positively selected only in periodontitis patients and negatively selected only in controls in the current dataset. In the dataset from study by Wang et al. [27], *pgpB* in *Porphyromonas gingivalis, macB* in *Candidatus Regiella insecticola* and *lnuC* from *Streptococcus agalactiae* all had a Ka/Ks rate > 1 in periodontitis patients and < 1 in controls. However, *macB* in *Candidatus Regiella insecticola* was found to be positively selected in our controls, but negatively selected in our periodontitis patients (Figure 5A). In the meantime, *lnuC* from *Streptococcus agalactiae* in the data by the previous study was positively selected both in our periodontitis patients and controls (Figure 5A and B). The *ileS* gene from *Bifidobacteria* was positively selected in both periodontitis patients and controls from both datasets, indicating a strong evolutionary dynamic (Figure 5A and B).

**Figure 5.**
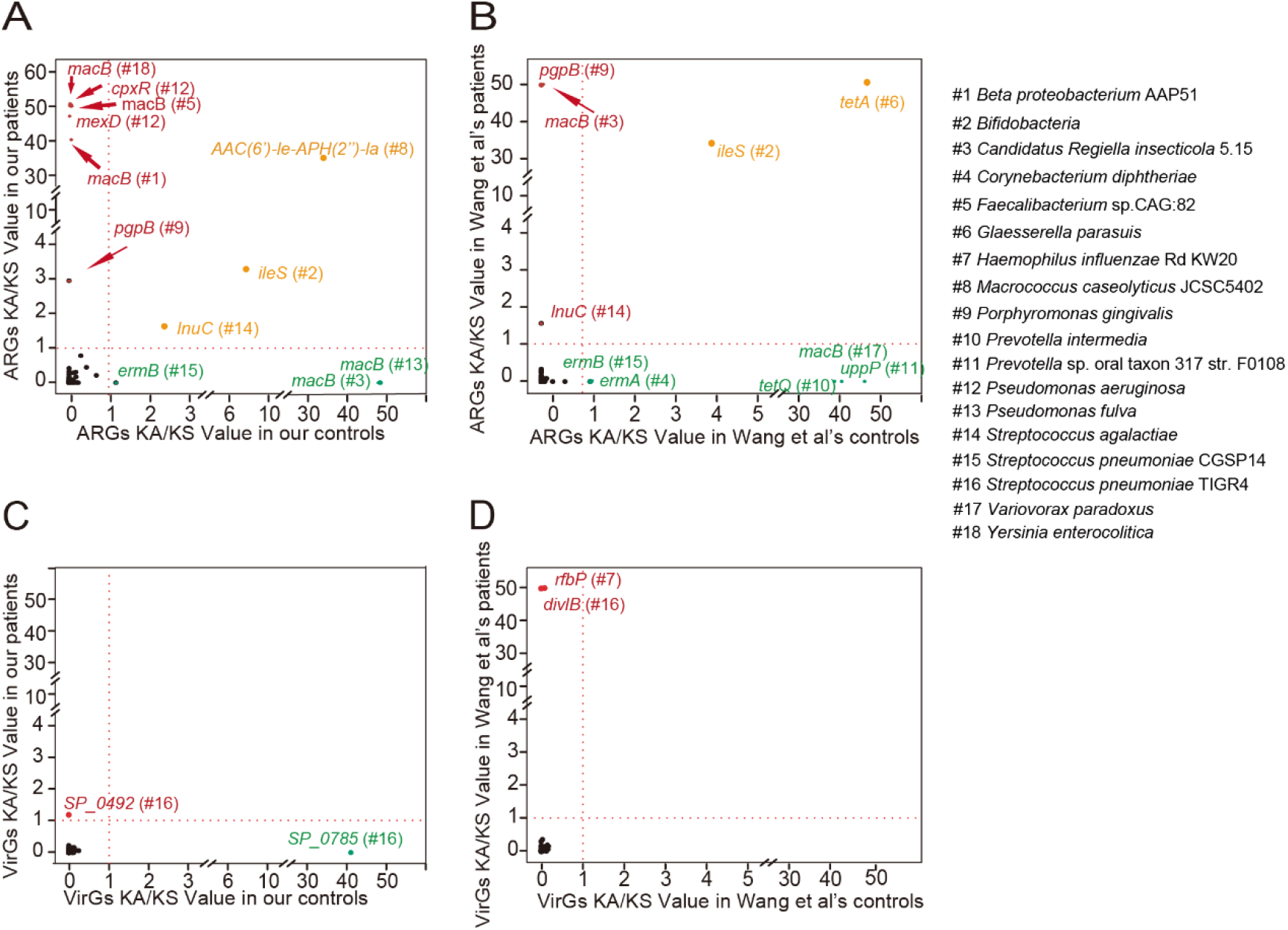
Ka/Ks values of ARGs and VirGs in periodontitis and control from both datasets. **A**. Ka/Ks values of ARGs in periodontitis and control from current dataset. **B**. Ka/Ks values of ARGs in periodontitis and control from Wang et al. 2016. **C**. Ka/Ks values of VirGs in periodontitis and control from current dataset. **D**. Ka/Ks values of VirGs in periodontitis and control from Wang et al. 2016. Genes are marked in red if their Ka/Ks values are higher than 1 in periodontitis samples and lower than 1 in control samples. Those marked in green are genes with only Ka/Ks values higher than 1 in control samples and lower than 1 in periodontitis samples. Genes are colored brown if their Ka/Ks values are higher than 1 in both periodontitis and control samples.

Besides ARGs, a total of 181 VirGs with at least one non-synonymous SNP were identified, which highlights the evolutionary potential in VirGs in both studies. Notably, 145 of these VirGs from pathogens were reported to be associated with respiratory infection (Table S13). We found a total of three VirGs positively selected only in periodontitis patients and negatively selected only in controls from both datasets (Figure 5C, D and Table S13). One gene from the current dataset was *SP_0492*, which encodes a conserved protein in *Streptococcus pneumoniae* TIGR4 (Figure 5C). The other two genes from the previous study were *divIB*, encoding a cell division protein in *Streptococcus pneumoniae* TIGR4, and r*fbP*, encoding an undecaprenyl-phosphate galactose phosphotransferase involved in biosynthesis of bacterial lipopolysaccharide in *Haemophilus influenzae* Rd KW20 (Figure 5D). Notably, all pathogens in which these VirGs were positively selected are associated with respiratory infection (Table S13).

## Discussion

Increased epidemiologic evidence supported the associations between periodontitis and systemic diseases, such as respiratory disease. Dysbiosis of the oral microflora lead to translocation of bacteria and bacterial metabolites to the airway, increasing risk of respiratory infection. In this study, we found a set of significantly abundant bacteria as well as functional pathways in periodontitis patients based on metagenomic sequence data from six periodontitis plaque specimens and three healthy periodontal plaque specimens. Through investigating potential bacteria, ARGs and VirGs associated with periodontitis, we found several positively selected genes encoding virulence proteins and antibiotic-resistance proteins encoded by respiratory disease-associated pathogens from both our dataset and the study by Wang et al. [27]. These findings indicate a potential association between periodontitis and respiratory infection.

Both Wilcoxon rank rum test and LEfSe analysis showed that two phyla and five species had significantly increased abundance in periodontitis patients than controls. *Spirochaetes* is one of the two phyla significantly increased in patients whose accumulation in subgingival plaque appears to be a marker of the clinical severity of periodontal disease [30], comprising up to 50% of the polymicrobial population in subgingival plaque in periodontitis, and < 1% in healthy periodontium [31]. Among members of *Spirochaetes, Treponema* is the only genus that appears in the human mouth [32], and which was also significantly enriched in our patients. Among species of *Treponema*, both *Treponema denticola* and *Treponema medium* were significantly enriched in our patients, and all were reported to be periodontitis pathogens [33]. *Treponema denticola* is an important pathogen associated with periodontitis, as it not only destroys host tissue but also co-aggregates with *Porphyromonas gingivalis, Fusobacterium nucleatum* and *Tannerella forsythia* to form the ‘red complex’ [34, 35]. *Synergistetes* is another significantly enriched phylum playing a role in increasing pocket depth, inflammation, anaerobiosis, and gingival tissue destruction [36]. Species members *Fretibacterium fastidiosum, Desulfobulbus* sp. oral taxon 041 and *Bacteroidetes bacterium* oral taxon 272, which were significantly enriched in our periodontitis patients, have been reported to be significantly associated with periodontitis [36-38].

Several EggNOG terms associated with drug-resistance and bacteria virulence were found to be significantly enriched in periodontitis patients. Transposase, the most frequently abundance of functional EggNOG terms, often harbor resistance genes against many different antibiotics. Most DNA transposons confer resistance to all major antibiotic classes against bacteria, and transpositions of these transposons are major drivers of resistance spreading [39, 40]. The significantly over-abundant cell wall/membrane/envelope biogenesis and MATE efflux family proteins in patients together facilitate drug effluxion and induce drug resistance [41, 42]. Active efflux could reduce the intracellular accumulation of antimicrobials, which is a major mechanism of antimicrobial resistance in pathogenic organisms [42]. It is crucially important to maintain an effective barrier around the organism for active efflux systems, to reduce the influx of antimicrobials into the cell [41]. GSEA analysis showed the enrichment for MCP and gingipain R in patients and genes associated with the synthesis of fatty acids in controls. MCPs, which are enriched in periodontitis patients, comprise a family of bacterial receptors that accept diverse environmental signals such as those from attractants and repellents and mediate the approach or escape through alteration of swimming behavior [43]. Gingipains are trypsin-like cysteine proteinases produced by *Porphyromonas gingivalis*, one of the most prominent virulence factors in periodontal disease. Gingipains not only deal with cell surface proteins when contacted but also degrade components of the cell-to-cell contacts and cause detachment of epithelial cells from the connective tissue of the gingiva [44, 45]. Several kind of genes associated with the synthesis of fatty acids, such as fatty acid synthase, esterase and MaoC domain protein dehydratase, were found to be enriched in controls, and some kinds of fatty acids have been reported to be of benefit for oral health and play a positive role in periodontitis treatment [46, 47].

We called SNPs and compared the MAF of non-synonymous and unique non-synonymous SNPs in ARGs between periodontitis patients and controls in both studies, showing that ARGs may suffer much more selection pressure in periodontitis patients compared with controls, which is similar to the findings by Zhao et al.[48]. Besides ARGs, we also found similar trends in VirGs, which indicated that VirGs may also be under more selection pressure in periodontitis patients compared with controls. In fact, inclusion criteria for subjects in our study were similar to the previous study by Wang et al. [27], including without systemic diseases and prosthetic dental appliances. In addition, all subjects had never received professional periodontal therapy or antibiotic treatment within the previous 3 months. Considering that the periodontitis and control subjects in the same study were all local residents and shared similar climate and environmental conditions without distinct dietary habits, the selection pressure may be associated with the interaction between pathogenesis and treatment.

The Ka/Ks ratio is commonly used in understanding the evolutionary dynamic of protein-coding sequences across closely related and yet divergent species [49-51]. Several ARGs were found with Ka/Ks > 1 in periodontitis patients and < 1 in controls. Proteins encoded by *cpxR* in *Pseudomonas aeruginosa* activate the expression of the MexAB-OprM efflux pump and enhance drug resistance [52]. *pgpB*, encoding lipid A 4’-phosphatase in *Porphyromonas gingivalis* ATCC 33277, conferred resistance to polymyxin B by making lipid A lose a negative charge from the lipid A 4’-phosphate [53]. It was noted that *Porphyromonas gingivalis* is one of the most import periodontitis pathogen and lipid A is one of the key virulence factors in *Porphyromonas gingivalis* invasion. *pgpB* was the only ARG existing in periodontitis-associated pathogens with Ka > Ks in periodontitis patient samples and Ka < Ks in controls; this gene was identical in both previous study by Wang et al. and our data [27]. Interestingly, *Porphyromonas gingivalis* could also cause pneumonia [12], and Polymyxin B resisted by PgpB has already been widely used in pneumonia treatment [54]. It was supposed that selection pressure of *pgpB* was likely to be associated with both periodontitis and pneumonia.

None of the Ka values of the virulence proteins in periodontitis pathogens were found to be higher than the Ks values in both datasets. However, three VirGs in respiratory tract pathogens were found to have Ka/Ks > 1 in periodontitis and Ka/Ks < 1 in controls, two of which were from *Streptococcus pneumoniae* TIGR4 and one that was involved in the biosynthesis of bacterial lipopolysaccharide in *Haemophilus influenzae* Rd KW20. When focusing on the background VirGs with at least one non-synonymous SNP, these three VirGs were positively selected. It was noted that nearly all (91.8%) of the background VirGs with at least one non-synonymous SNP for natural selection were from respiratory tract pathogens. The evolutionary background of VirGs in periodontitis may suggest a possible association with respiratory infection.

Multiple epidemiological studies have reported an association between periodontal inflammation and respiratory infection [55-59]. Poor oral hygiene and periodontal disease may facilitate the colonization of potential respiratory pathogens (PRPs) [60], especially on the surface of dental plaque. Once the host’s defense is compromised, PRPs can access the respiratory tract by oropharyngeal secretions or inhalation, and then overwhelm the host’s immune system [11]. Some pathogens associated with periodontitis, such as *Porphyromonas gingivalis* and *Treponema denticola*, are also reported to cause pneumonia [12]. It may be hypothesized that in our study, although the pathogens associated with respiratory infection were not significantly dominant in periodontitis, some of the virulence proteins or the antibiotic resistance proteins of these pathogens were significantly abundant and putatively positively selected; therefore, these pathogens could have been ready to access and invade the respiratory tract. It is likely that there could be an interaction between these genes and respiratory disease in periodontitis.

## Conclusion

Although the oral microbial profile of periodontitis is still unclear, the present study provides several points of clarification. Apart from increased abundance of pathogens and community diversity consistent with previously reported studies, we found a new pathogen and several abundant functional pathways that differed between periodontitis patients and healthy controls. We also found that ARGs as well as VirGs may also be subject to positive selection. Several identical significantly abundant and positively selected ARGs and VirGs were identified in respiratory disease-associated pathogens in the two different datasets. These genes could not only serve as targets for further studies but may also offer clues for studying the association between respiratory disease and periodontitis. Our findings may enrich the knowledge of pathogens, drug resistance and functional pathways in the oral microbiota of periodontitis and shed lights on the association of periodontitis with respiratory disease. However, animal model or deep functional assays should be carried out to explore the functional mechanisms of pathogens in periodontitis and the association between periodontal inflammation and systemic diseases in the future.

## Materials and methods

### Sample and data collection

A total of 30 periodontitis patients and 15 periodontal healthy individuals were recruited based on the defined standard description in a previous study [61]. Participants were all previously un-treated for periodontitis. Periodontal health was defined as having a probing depth and attachment loss < 2 mm. Participants with any other prosthetic dental appliances, antibiotic treatment within the previous 3 months, diabetes, hypertension or hepatitis were excluded. Clinical details for each participant are provided in the Table S1. The periodontist performed a full-mouth examination and took dental plaque samples from the tooth with the poorest condition, excluding third molars.

For each individual, dental plaque was collected from 3-4 sites (buccal surface, lingual surface and supragingival sites) by a periodontist and pooled together as one sample. Dental plaque was collected using a sterile inoculating-loop-like device to increase accumulation and to avoid gingival bleeding. Each plaque sample was placed in a sterile 1.5 ml centrifuge tube containing 50 μl of phosphate buffer solution (PBS; pH = 8.0) and stored at −80 °C until use. All specimens were collected in a sterilized operating room with strict aseptic operation.

Previous raw sequencing data for dental plaque (PHP1-10, PDP1-10) from the study by Wang et al. was downloaded from the NCBI SRA database with accession number SRP033553 [27]. The ARGs and VirGs were analyzed with SRA toolkit V2.10.0 with the “prefetch” command. The “fastq-dump” command was used to obtain the FASTQ files from the downloaded SRA file.

### Specimen processing and metagenomic sequencing

Metagenomic DNA was isolated from the 45 specimens individually using a QIAamp DNA Mini Kit (51306, Qiagen, Hilden, Germany). The quality and quantity of isolated DNA was measured using a Qubit Fluorometer, a Nano Drop 2000 spectrophotometer (Thermo Fisher Scientific, Waltham, MA, USA), and agarose gel electrophoresis. For each specimen, 0.5 μg of purified metagenomic DNA was sheared into fragments of ∼180 bp in length, which then were end-polished, A-tailed, and ligated with the full-length adaptor for library construction according to a standard protocol provided by Illumina, Inc. (San Diego, CA, USA). After further PCR amplification, the PCR products were purified using the AMPure XP system (A63880, Beckman Coulter, Brea, CA). Finally, the libraries were analyzed for size distribution with the Agilent2100 bioanalyzer (Agilent Technologies, Palo Alto, CA) and quantified using real-time PCR before high-throughput sequencing on an Illumina HiSeq 2500.

### Read filtering and merging

First, the quality control, cutoff of the adapter, and removal of the low-quality reads were performed as described previously[62]. Second, filtered high-quality PE reads were aligned to human genome assembly (hg19) using Bowtie2 v2.3.4 to remove human contamination, and further visualized through Splicing Viewer [63]. Finally, the nonhuman PE reads were merged using FLASH v1.2.11 with overlap lengths ranging from 6 to 40 bp and mismatch thresholds set to 0.15. These filtering and merging steps were processed for the current dataset and the dataset from Wang et al. [27].

### Taxonomic classification, construction of a nonredundant reference gene set and functional annotation

MetaPhlAnv2.7.7 [24] was used to process the taxonomic profiles, and the Wilcoxon-rank sum test was used to determine the differentially abundance of microbial taxa between periodontitis patients and controls. Based on the phylum and species abundance profile, the beta diversity was estimated by PCA and NMDS analysis, and the alpha diversity was measured using the Shannon index, Simpson index, InverseSimpson index, and Pielou index. LEfSe [64] analysis was performed to obtain taxonomic biomarkers. To obtain a non-redundant reference gene set, all FASTA files were integrated, and then MEGAHIT v1.1.3 was used to assemble these reads into contigs, in which only contigs with a length of 500 bp or more were retained. A set of proteins predicted by MetaGeneMark v3.8 was input into CD-HIT v4.6.2 to obtain the nonredundant reference protein sequence set, and a nonredundant nucleotide reference gene set was also obtained from the predicted nucleotide sequences according to the gene number. Reads were aligned to the nonredundant nucleotide gene set using Bowtie2, and the gene RPK value was calculated.

EggNOG-mapper v1.0.3 [65] was used to map the correlated protein sequences of all predicted genes to annotation databases, such as KEGG, EggNOG, and GO. The RPK value of each annotation was obtained by summing the RPK values of genes with the same annotation, and DESeq2 was used to obtain the significant functional annotations. GSEA was performed using GSEA 3.0 with RPK values in genes from both patients and controls as well as genes included in KEGG terms, GO terms and EggNOG terms.

### ARG and VirG annotation

The abundances of each gene family in the oral microbiota were analyzed by HUMAnN2 v0.11.1 [66] in RPK units. The significantly enriched gene families were obtained using DESeq2. DeepARG [26] was used to annotate all ARG reads in each sample in our data and the data by Wang et al. [27]. The DIAMOND cutoff was set to 50% for the threshold of sequence matching ratio as well as 60% for similarity and 1.00×10^−7^ for the e-value as recommended by ARGs-OAP [67]. The relative abundance of each ARG was calculated as described previously [68]; and LEfSe [64] analysis was used to obtain the ARG biomarkers. Virulence protein sequences were downloaded from PATRIC [69], Victors [70] and the virulence factor database (VFDB) [71], and a nonredundant reference virulence sequence was finally obtained after using CD-HIT. All reads in each sample in our data and the data from the previous study were annotated using the same method and cutoff values as those in the ARG annotation.

### Reference ARG and VirG nucleotide sequence construction, SNP calling and positively selected gene detection

The reference ARG protein sequences processed in DeepARG were used to construct a custom database. Among a total of 14,974 nonredundant ARG proteins, there were 10,619 protein sequences from UniProt90, 2,152 from the Comprehensive Antibiotic Research Database (CARD), and 2,203 from the Antibiotic Resistance Genes Database (ARDB). Almost all correlated nucleotide sequences were downloaded from the official website. To ensure the accuracy, the downloaded nucleotide sequences were aligned against the reference protein sequences using BLASTX, and the matched nucleotide sequence segment in the best hit in each blast was considered as the correlating reference nucleotide sequence.

The FASTA files of each sample were aligned to the constructed reference nucleotide sequences using BWA-mem v0.7.17. SAMtools v1.9, and BCFtools v1.9. VarScan v2.4.3 were used to call SNPs and VCFtools v0.1.16 was used to filter the low-quality SNPs from each sample[72]. SNPs called by BCFtools were combined with those called by VarScan. According to the Bacterial and Plant Plastid Code, the amino acid coded by an altered codon was compared with that coded by a reference codon to determine whether the SNP was a synonymous or non-synonymous SNP. The SNP rarefaction curve was plotted using the R package “Vegan”, and the Ka/Ks rate was calculated using the KaKs calculator after producing the axt file [48].

The non-redundant reference virulence protein sequences were aligned to the NT database downloaded from NCBI using TBLASTN. The aligned nucleic acid sequence with the best hit score was considered the corresponding nucleic acid sequence. The subsequent SNP calling and annotation methods were similar to those used in the calling and annotation of ARG SNPs.

## Data Availability

The data that support the findings of this study have been deposited in the Genome Sequence Archive in National Genomics Data Center, Beijing Institute of Genomics (China National Center for Bioinformation), Chinese Academy of Sciences, under accession number CRA003553 that are publicly accessible at https://bigd.big.ac.cn/gsa.

## Ethical statements

This study was approved by the research institute’s institutional review board, and written informed consent was received from all participants before their enrolment.

## Data availability

The raw sequence data reported in this paper have been deposited in the Genome Sequence Archive [73] in National Genomics Data Center [74], Beijing Institute of Genomics (China National Center for Bioinformation), Chinese Academy of Sciences, under accession number **CRA003553** that are publicly accessible at https://bigd.big.ac.cn/gsa.

## CRediT author statement

**Zhenwei Liu:** Formal analysis, Visualization, Writing-Original Draft, Writing-Review & Editing. **Tao Zhang:** Methodology, Formal analysis, Visualization, Writing-Original Draft. **Keke Wu:** Software, Data Curation, Visualization, Writing-Original Draft. **Zhongshan Li:** Formal analysis, Data Curation. **Xiaomin Chen:** Investigation, Resources. **kairui Qiu:** Data Curation, Investigation. **Shan Jiang:** Formal analysis, Validation. **Lifeng Du:** Data Curation, Resources. **Saisai Lu:** Investigation, Validation. **Chongxiang Lin:** Resources, Data Curation. **Jinyu Wu:** Conceptualization, Writing-Original draft, Writing-Review & Editing, Supervision. **Xiaobing Wang:** Conceptualization, Writing-Original draft, Writing-Review & Editing, Supervision, Funding acquisition. All authors read and approved the final manuscript.

## Acknowledgments

This study was supported by the National Natural Science Foundation of China (Grant No. 81700062), Science and Technology Project of Zhejiang Provincial Health Commission (Grant No. 2019RC050) and the Science and Technology Project of Wenzhou (Grant No. Y20160028). We thank Helen Jeays, BDSc AE, from Liwen Bianji, Edanz Editing China (www.liwenbianji.cn/ac), for editing the English text of a draft of this manuscript. We also thank Jianmin Wu for providing insightful comments and suggestions on this manuscript.

## Supplementary material

**Figure S1 Quality control of raw and clean reads**

**A**. Clean, raw and uncontaminated read numbers as well as the clean reads rate and non-human contaminated reads rate of our data. **B**. Read number and rates of NS reads (reads with N bases), low-quality reads, and polluted adapter reads in current dataset.

**Figure S2 The shift of oral microbiota between periodontitis and control samples at the genus level**

**A**. The difference in relative abundance of each genus between periodontitis and control samples among the top 30 genera. The significantly differential relative abundance for genera was marked by red (alpha = 0.05, Wilcoxon rank sum test). **B**. Rarefaction curve at the species level. **C**. Genus biomarkers in periodontitis and control samples were depicted by LEfSe analysis (LDA > 3).

**Figure S3 Beta diversity and alpha diversity at the phylum or species level**

**A**. PCA analysis at the phylum level. **B**. Non-metric multidimensional scaling (NMDS) analysis at the phylum level. **C**. ANOSIM of NMDS analysis results at the phylum level.

**D**. InverseSimpson index at the phylum level between periodontitis and control samples.

**E**. Pielou index at the phylum level between periodontitis and control samples. **F**. Shannon index at the phylum level between periodontitis and control samples. **G**. Simpson index at the phylum level between periodontitis and control samples. **H**. Difference in InverseSimpson index at the species level between periodontitis and control samples. **I**. Simpson index at the species level between periodontitis and control samples. All *P* values were calculated by Wilcoxon rank sum test.

**Figure S4 Comparison of the relative abundance of UniProt90 genes between the both datasets**

**A**. Principal component analysis (PCA) analysis between two datasets. **B**. The non-metric multidimensional scaling (NMDS) analysis of all samples using UniProt90 gene relative abundance from both datasets. **C**. Intersections between antibiotic resistance genes (ARGs) in our patient samples and Wang’s patient samples. **D**. Intersections between virulence genes (VirGs) in our patient samples and Wang’s patient samples.

**Figure S5 Rarefaction curve of SNPs in ARGs and VirGs from both datasets**

Rarefaction curve of SNPs in ARGs (**A**) and VirGs (**B**) from the current dataset. (c, d) Rarefaction curve of SNPs in ARGs (**C**) and VirGs (**D**) of periodontitis patients from Wang et al. 2016. Rarefaction curve of SNPs in ARGs (**E**) and VirGs (**F**) of controls from Wang et al. 2016. SNPs, single-nucleotide polymorphisms.

**Figure S6 Comparison of synonymous SNPs in ARGs and VirGs between periodontitis and control samples from both datasets**

Distribution of the minor allele frequency (MAF) of SNPs in ARGs from both current study (**A**) and previous study (**B**). **C**. Comparison of the MAF of synonymous SNPs in ARGs between periodontitis patients and controls in both datasets. Distribution of the MAF of synonymous SNPs in VirGs both current study (**D**) and previous study (**E**). **F**. Comparison of the MAF of synonymous SNPs in VirGs between periodontitis patients and controls in both datasets. All *P* values were calculated by Wilcoxon rank sum test.

**Table S1**. Clinical information for all participants.

**Table S2**. Quality control of our sequenced data.

**Table S3**. Relative abundance of kingdom level in the current samples.

**Table S4**. Relative abundance of phylum level in the current samples.

**Table S5**. Relative abundance of genus level in the current samples.

**Table S6**. Relative abundance of species level in the current samples.

**Table S7**. The differentially function of microbial taxa between periodontitis patients and controls.

**Table S8**. The GSEA analysis in KEGG, GO, EggNOG terms.

**Table S9**. LEfSe analysis of antibiotic resistance genes (ARGs) and virulence genes (Virs) in the current dataset.

**Table S10**. Quality control of dataset by Wang et al.2016.

**Table S11**. LEfSe analysis of antibiotic resistance genes (ARGs) and virulence genes (Virs) in the dataset by Wang et al. 2016.

**Table S12**. The Ka/Ks values of each ARG in both datasets.

**Table S13**. The Ka/Ks values of each VirGs in both datasets.

## REFERENCE

[1] Darveau RP. Periodontitis: A polymicrobial disruption of host homeostasis. Nat Rev Microbiol 2010;8:481–90.

[2] Hernandez M, Dutzan N, Garcia-Sesnich J, Abusleme L, Dezerega A, Silva N, et al. Host-pathogen interactions in progressive chronic periodontitis. J Dent Res 2011;90:1164–70.

[3] Jiao Y, Hasegawa M, Inohara N. The role of oral pathobionts in dysbiosis during periodontitis development. J Dent Res 2014;93:539–46.

[4] Chukkapalli SS, Easwaran M, Rivera-Kweh MF, Velsko IM, Ambadapadi S, Dai J, et al. Sequential colonization of periodontal pathogens in induction of periodontal disease and atherosclerosis in ldlrnull mice. Pathog Dis 2017;75.

[5] Liccardo D, Cannavo A, Spagnuolo G, Ferrara N, Cittadini A, Rengo C, et al. Periodontal disease: A risk factor for diabetes and cardiovascular disease. Int J Mol Sci 2019;20.

[6] Cobb CM, Kelly PJ, Williams KB, Babbar S, Angolkar M, Derman RJ. The oral microbiome and adverse pregnancy outcomes. Int J Womens Health 2017;9:551–9.

[7] Ide M, Harris M, Stevens A, Sussams R, Hopkins V, Culliford D, et al. Periodontitis and cognitive decline in Alzheimer’s disease. PLoS One 2016;11:e0151081.

[8] Potempa J, Mydel P, Koziel J. The case for periodontitis in the pathogenesis of rheumatoid arthritis. Nat Rev Rheumatol 2017;13:606–20.

[9] Flemer B, Warren RD, Barrett MP, Cisek K, Das A, Jeffery IB, et al. The oral microbiota in colorectal cancer is distinctive and predictive. Gut 2018;67:1454–63.

[10] de Molon RS, Rossa C, Jr., Thurlings RM, Cirelli JA, Koenders MI. Linkage of periodontitis and rheumatoid arthritis: Current evidence and potential biological interactions. Int J Mol Sci 2019;20.

[11] Bansal M, Khatri M, Taneja V. Potential role of periodontal infection in respiratory diseases – a review. J Med Life 2013;6:244–8.

[12] Kimizuka R, Kato T, Ishihara K, Okuda K. Mixed infections with Porphyromonas gingivalis and Treponema denticola cause excessive inflammatory responses in a mouse pneumonia model compared with monoinfections. Microbes Infect 2003;5:1357–62.

[13] Costalonga M, Herzberg MC. The oral microbiome and the immunobiology of periodontal disease and caries. Immunol Lett 2014;162:22–38.

[14] Roy S, Douglas CW, Stafford GP. A novel sialic acid utilization and uptake system in the periodontal pathogen Tannerella forsythia. J Bacteriol 2010;192:2285–93.

[15] Sudhakara P, Sellamuthu I, Aruni AW. Bacterial sialoglycosidases in virulence and pathogenesis. Pathogens 2019;8.

[16] Curtis MA, Diaz PI, Van Dyke TE. The role of the microbiota in periodontal disease. Periodontol 2000 2020;83:14–25.

[17] Solbiati J, Frias-Lopez J. Metatranscriptome of the oral microbiome in health and disease. J Dent Res 2018;97:492–500.

[18] Read AF, Woods RJ. Antibiotic resistance management. Evol Med Public Health 2014;2014:147.

[19] Ventola CL. The antibiotic resistance crisis: Part 1: Causes and threats. P T 2015;40:277–83.

[20] zur Wiesch PA, Kouyos R, Engelstadter J, Regoes RR, Bonhoeffer S. Population biological principles of drug-resistance evolution in infectious diseases. Lancet Infect Dis 2011;11:236–47.

[21] Rams TE, Degener JE, van Winkelhoff AJ. Antibiotic resistance in human chronic periodontitis microbiota. J Periodontol 2014;85:160–9.

[22] Ardila CM, Granada MI, Guzman IC. Antibiotic resistance of subgingival species in chronic periodontitis patients. J Periodontal Res 2010;45:557–63.

[23] van Winkelhoff AJ, Herrera D, Oteo A, Sanz M. Antimicrobial profiles of periodontal pathogens isolated from periodontitis patients in the Netherlands and Spain. J Clin Periodontol 2005;32:893–8.

[24] Truong DT, Franzosa EA, Tickle TL, Scholz M, Weingart G, Pasolli E, et al. Metaphlan2 for enhanced metagenomic taxonomic profiling. Nat Methods 2015;12:902–3.

[25] Nikaido H. Multiple antibiotic resistance and efflux. Curr Opin Microbiol 1998;1:516–23.

[26] Arango-Argoty G, Garner E, Pruden A, Heath LS, Vikesland P, Zhang L. Deeparg: A deep learning approach for predicting antibiotic resistance genes from metagenomic data. Microbiome 2018;6:23.

[27] Wang J, Gao Y, Zhao F. Phage-bacteria interaction network in human oral microbiome. Environ Microbiol 2016;18:2143–58.

[28] Wang DP, Wan HL, Zhang S, Yu J. Gamma-MYN: A new algorithm for estimating ka and ks with consideration of variable substitution rates. Biol Direct 2009;4:20.

[29] Liu G, Wu J, Yang H, Bao Q. Codon usage patterns in Corynebacterium glutamicum: Mutational bias, natural selection and amino acid conservation. Comp Funct Genomics 2010;2010:343569.

[30] Loesche WJ. The role of spirochetes in periodontal disease. Adv Dent Res 1988;2:275–83.

[31] Chan EC, McLaughlin R. Taxonomy and virulence of oral spirochetes. Oral Microbiol Immunol 2000;15:1–9.

[32] Dewhirst FE, Chen T, Izard J, Paster BJ, Tanner AC, Yu WH, et al. The human oral microbiome. J Bacteriol 2010;192:5002–17.

[33] Belda-Ferre P, Alcaraz LD, Cabrera-Rubio R, Romero H, Simon-Soro A, Pignatelli M, et al. The oral metagenome in health and disease. ISME J 2012;6:46–56.

[34] Ikegami A, Honma K, Sharma A, Kuramitsu HK. Multiple functions of the leucine-rich repeat protein lrra of Treponema denticola. Infect Immun 2004;72:4619–27.

[35] Rosen G, Genzler T, Sela MN. Coaggregation of Treponema denticola with Porphyromonas gingivalis and Fusobacterium nucleatum is mediated by the major outer sheath protein of Treponema denticola. FEMS Microbiol Lett 2008;289:59–66.

[36] You M, Mo S, Watt RM, Leung WK. Prevalence and diversity of Synergistetes taxa in periodontal health and disease. J Periodontal Res 2013;48:159–68.

[37] Gao W, Chan Y, You M, Lacap-Bugler DC, Leung WK, Watt RM. In-depth snapshot of the equine subgingival microbiome. Microb Pathog 2016;94:76–89.

[38] Cao Y, Qiao M, Tian Z, Yu Y, Xu B, Lao W, et al. Comparative analyses of subgingival microbiome in chronic periodontitis patients with and without IgA nephropathy by high throughput 16S rRNA sequencing. Cell Physiol Biochem 2018;47:774–83.

[39] Lovell S, Goryshin IY, Reznikoff WR, Rayment I. Two-metal active site binding of a Tn5 transposase synaptic complex. Nat Struct Biol 2002;9:278–81.

[40] Rubio-Cosials A, Schulz EC, Lambertsen L, Smyshlyaev G, Rojas-Cordova C, Forslund K, et al. Transposase-DNA complex structures reveal mechanisms for conjugative transposition of antibiotic resistance. Cell 2018;173:208–20 e20.

[41] Nikaido H. Preventing drug access to targets: cell surface permeability barriers and active efflux in bacteria. Semin Cell Dev Biol 2001;12:215–23.

[42] Borges-Walmsley MI, Walmsley AR. The structure and function of drug pumps. Trends Microbiol 2001;9:71–9.

[43] Derr P, Boder E, Goulian M. Changing the specificity of a bacterial chemoreceptor. J Mol Biol 2006;355:923–32.

[44] Nakayama M, Ohara N. Molecular mechanisms of Porphyromonas gingivalis-host cell interaction on periodontal diseases. Jpn Dent Sci Rev 2017;53:134–40.

[45] Sheets SM, Robles-Price AG, McKenzie RM, Casiano CA, Fletcher HM. Gingipain-dependent interactions with the host are important for survival of Porphyromonas gingivalis. Front Biosci 2008;13:3215–38.

[46] Kesavalu L, Bakthavatchalu V, Rahman MM, Su J, Raghu B, Dawson D, et al. Omega-3 fatty acid regulates inflammatory cytokine/mediator messenger RNA expression in Porphyromonas gingivalis-induced experimental periodontal disease. Oral Microbiol Immunol 2007;22:232–9.

[47] Sommakia S, Baker OJ. Regulation of inflammation by lipid mediators in oral diseases. Oral Dis 2017;23:576–97.

[48] Zhao F, Bai J, Wu J, Liu J, Zhou M, Xia S, et al. Sequencing and genetic variation of multidrug resistance plasmids in Klebsiella pneumoniae. PLoS One 2010;5:e10141.

[49] Yu T, Li J, Yang Y, Qi L, Chen B, Zhao F, et al. Codon usage patterns and adaptive evolution of marine unicellular cyanobacteria Synechococcus and Prochlorococcus. Mol Phylogenet Evol 2012;62:206–13.

[50] Zhang Z, Li J, Zhao XQ, Wang J, Wong GK, Yu J. Kaks_calculator: Calculating ka and ks through model selection and model averaging. Genomics Proteomics Bioinformatics 2006;4:259–63.

[51] Li WH. Unbiased estimation of the rates of synonymous and nonsynonymous substitution. J Mol Evol 1993;36:96–9.

[52] Tian ZX, Yi XX, Cho A, O’Gara F, Wang YP. CpxR activates MexAB-oprm efflux pump expression and enhances antibiotic resistance in both laboratory and clinical nalB-type isolates of Pseudomonas aeruginosa. PLoS Pathog 2016;12:e1005932.

[53] Coats SR, To TT, Jain S, Braham PH, Darveau RP. Porphyromonas gingivalis resistance to polymyxin B is determined by the lipid A 4’-phosphatase, PGN_0524. Int J Oral Sci 2009;1:126–35.

[54] Palmer LB. Aerosolized antibiotics in the intensive care unit. Clin Chest Med 2011;32:559–74.

[55] Sjogren P, Nilsson E, Forsell M, Johansson O, Hoogstraate J. A systematic review of the preventive effect of oral hygiene on pneumonia and respiratory tract infection in elderly people in hospitals and nursing homes: Effect estimates and methodological quality of randomized controlled trials. J Am Geriatr Soc 2008;56:2124–30.

[56] Scannapieco FA, Ho AW. Potential associations between chronic respiratory disease and periodontal disease: Analysis of National Health and Nutrition Examination Survey III. J Periodontol 2001;72:50–6.

[57] Scannapieco FA. Role of oral bacteria in respiratory infection. J Periodontol 1999;70:793–802.

[58] Paju S, Scannapieco FA. Oral biofilms, periodontitis, and pulmonary infections. Oral Dis 2007;13:508–12.

[59] Laurence B, Mould-Millman NK, Scannapieco FA, Abron A. Hospital admissions for pneumonia more likely with concomitant dental infections. Clin Oral Investig 2015;19:1261–8.

[60] Scannapieco FA, Mylotte JM. Relationships between periodontal disease and bacterial pneumonia. J Periodontol 1996;67:1114–22.

[61] Wang J, Qi J, Zhao H, He S, Zhang Y, Wei S, et al. Metagenomic sequencing reveals microbiota and its functional potential associated with periodontal disease. Sci Rep 2013;3:1843.

[62] Wang T, Liu Q, Li X, Wang X, Li J, Zhu X, et al. RRBS-Analyser: A comprehensive web server for reduced representation bisulfite sequencing data analysis. Hum Mutat 2013;34:1606–10.

[63] Liu Q, Chen C, Shen E, Zhao F, Sun Z, Wu J. Detection, annotation and visualization of alternative splicing from RNA-Seq data with SplicingViewer. Genomics 2012;99:178–82.

[64] Segata N, Izard J, Waldron L, Gevers D, Miropolsky L, Garrett WS, et al. Metagenomic biomarker discovery and explanation. Genome Biol 2011;12:R60.

[65] Huerta-Cepas J, Forslund K, Coelho LP, Szklarczyk D, Jensen LJ, von Mering C, et al. Fast genome-wide functional annotation through orthology assignment by eggNOG-Mapper. Mol Biol Evol 2017;34:2115–22.

[66] Franzosa EA, McIver LJ, Rahnavard G, Thompson LR, Schirmer M, Weingart G, et al. Species-level functional profiling of metagenomes and metatranscriptomes. Nat Methods 2018;15:962–8.

[67] Yang Y, Jiang X, Chai B, Ma L, Li B, Zhang A, et al. ARGs-OAP: Online analysis pipeline for antibiotic resistance genes detection from metagenomic data using an integrated structured ARG-database. Bioinformatics 2016;32:2346–51.

[68] Hu Y, Yang X, Qin J, Lu N, Cheng G, Wu N, et al. Metagenome-wide analysis of antibiotic resistance genes in a large cohort of human gut microbiota. Nat Commun 2013;4:2151.

[69] Driscoll T, Gabbard JL, Mao C, Dalay O, Shukla M, Freifeld CC, et al. Integration and visualization of host-pathogen data related to infectious diseases. Bioinformatics 2011;27:2279–87.

[70] Sayers S, Li L, Ong E, Deng S, Fu G, Lin Y, et al. Victors: A web-based knowledge base of virulence factors in human and animal pathogens. Nucleic Acids Res 2019;47:D693–D700.

[71] 1 Liu B, Zheng D, Jin Q, Chen L, Yang J. VFDB 2019: A comparative pathogenomic platform with an interactive web interface. Nucleic Acids Res 2019;47:D687–D92.

[72] Chen Y, Li Z, Hu S, Zhang J, Wu J, Shao N, et al. Gut metagenomes of type 2 diabetic patients have characteristic single-nucleotide polymorphism distribution in Bacteroides coprocola. Microbiome 2017;5:15.

[73] Wang Y, Song F, Zhu J, Zhang S, Yang Y, Chen T, et al. GSA: Genome Sequence Archive. Genomics Proteomics Bioinformatics 2017;15:14–8.

[74] National Genomics Data Center M, Partners. Database resources of the National Genomics Data Center in 2020. Nucleic Acids Res 2020;48:D24–D33.

