## Supplementary material for "Metagenomic Analysis Reveals A Possible Association Between Respiratory Infection and Periodontitis": Table S2.docx

**Table S2. Quality control of our sequenced data.**

| **Sample** | **C1** | **C2** | **C3** | **P1** | **P2** | **P3** | **P4** | **P5** | **P6** |
| --- | --- | --- | --- | --- | --- | --- | --- | --- | --- |
| Raw read number | 31,581,072 | 32,031,842 | 31,934,538 | 29,952,888 | 31,985,284 | 32,282,950 | 31,413,656 | 31,755,984 | 31,185,194 |
| Raw base number | 4,737,160,800 | 4,804,776,300 | 4,790,180,700 | 4,492,933,200 | 4,797,792,600 | 4,842,442,500 | 4,712,048,400 | 4,763,397,600 | 4,677,779,100 |
| Clean read number | 30,407,662 | 30,834,978 | 30,748,896 | 28,753,246 | 30,912,934 | 30,825,240 | 29,988,486 | 30,633,470 | 29,782,530 |
| Clean read rate (%) | 96.28 | 96.26 | 96.29 | 96 | 96.65 | 95.48 | 95.46 | 96.47 | 95.5 |
| Clean base number | 4,561,149,300 | 4,625,246,700 | 4,612,334,400 | 4,312,986,900 | 4,636,940,100 | 4,623,786,000 | 4,498,272,900 | 4,595,020,500 | 4,467,379,500 |
| Low-quality read number | 247,298 | 260,812 | 213,588 | 239,604 | 326,804 | 234,942 | 227,160 | 234,800 | 200,644 |
| Low-quality read rate (%) | 0.78 | 0.81 | 0.67 | 0.8 | 1.02 | 0.73 | 0.72 | 0.74 | 0.64 |
| Ns read number | 38 | 42 | 32 | 22 | 36 | 30 | 38 | 28 | 40 |
| Ns read rate (%) | 0 | 0 | 0 | 0 | 0 | 0 | 0 | 0 | 0 |
| Raw Q30 base rate (%) | 92.16 | 92.2 | 92.68 | 92.21 | 92.04 | 92.84 | 92.56 | 92.51 | 92.92 |
| Clean Q30 base rate (%) | 92.48 | 92.53 | 92.97 | 92.54 | 92.47 | 93.14 | 92.85 | 92.81 | 93.17 |
| Clean read number | 30,407,662 | 30,834,978 | 30,748,896 | 28,753,246 | 30,912,934 | 30,825,240 | 29,988,486 | 30,633,470 | 29,782,530 |
| Reads after removal of  human sequence | 6,775,181 | 5,470,215 | 6,979,444 | 4,311,742 | 7,890,277 | 6,319,918 | 6,228,684 | 3,602,135 | 3,580,990 |
| Human sequence read rate (%) | 77.72 | 82.26 | 77.3 | 85 | 74.48 | 79.5 | 79.23 | 88.24 | 87.98 |
| Non-human sequence read Rate (%) | 22.28 | 17.74 | 22.7 | 15 | 25.52 | 20.5 | 20.77 | 11.76 | 12.02 |
