## Supplementary material for "Metagenomic Analysis Reveals A Possible Association Between Respiratory Infection and Periodontitis": Table S3.docx

**Table S3. Relative abundance of kingdom level in the current samples.**

| **Kingdom** | **C1** | **C2** | **C3** | **P1** | **P2** | **P3** | **P4** | **P5** | **P6** |
| --- | --- | --- | --- | --- | --- | --- | --- | --- | --- |
| Archaea | 0.000 | 0.000 | 0.000 | 0.000 | 0.000 | 0.055 | 0.000 | 0.000 | 0.000 |
| Bacteria | 100.000 | 100.000 | 100.000 | 99.845 | 100.000 | 99.945 | 100.000 | 100.000 | 100.000 |
| Viruses | 0.000 | 0.000 | 0.000 | 0.155 | 0.000 | 0.000 | 0.000 | 0.000 | 0.000 |
