## Supplementary material for "Metagenomic Analysis Reveals A Possible Association Between Respiratory Infection and Periodontitis": Table S4.docx

**Table S4. Relative abundance of phylum level in the current samples.**

| **Phylum** | **C1** | **C2** | **C3** | **P1** | **P2** | **P3** | **P4** | **P5** | **P6** | **Median C** | **Median P** | ***P* value** | **FDR** |
| --- | --- | --- | --- | --- | --- | --- | --- | --- | --- | --- | --- | --- | --- |
| Fusobacteria | 8.049 | 11.157 | 12.8 | 10.341 | 3.777 | 7.539 | 3.721 | 8.395 | 5.86 | 11.157 | 6.6995 | 0.095 | 0.762 |
| Actinobacteria | 16.15 | 11.052 | 10.898 | 3.854 | 3.625 | 6.709 | 12.922 | 8.152 | 5.032 | 11.052 | 5.8705 | 0.095 | 0.762 |
| Candidatus_  Saccharibacteria | 2.426 | 6.583 | 3.124 | 3.756 | 0.328 | 4.727 | 2.125 | 1.033 | 3.525 | 3.124 | 2.825 | 0.548 | 1.000 |
| Proteobacteria | 6.876 | 15.525 | 13.778 | 15.495 | 10.348 | 6.609 | 20.589 | 10.798 | 6.382 | 13.778 | 10.573 | 0.714 | 1.000 |
| Firmicutes | 17.146 | 14.608 | 19.878 | 20.279 | 20.049 | 17.846 | 11.426 | 17.52 | 14.553 | 17.146 | 17.683 | 0.905 | 1.000 |
| Euryarchaeota | 0 | 0 | 0 | 0 | 0 | 0.055 | 0 | 0 | 0 | 0 | 0 | 0.637 | 1.000 |
| Viruses_noname | 0 | 0 | 0 | 0.155 | 0 | 0 | 0 | 0 | 0 | 0 | 0 | 0.637 | 1.000 |
| Bacteroidetes | 44.334 | 36.731 | 37.567 | 35.091 | 52.248 | 47.211 | 40.361 | 48.634 | 59.403 | 37.567 | 47.9225 | 0.262 | 1.000 |
| Spirochaetes | 4.899 | 4.277 | 1.912 | 9.515 | 8.932 | 9.021 | 7.961 | 5.256 | 5.033 | 4.277 | 8.4465 | 0.024 | 0.238 |
| Synergistetes | 0.12 | 0.068 | 0.043 | 1.513 | 0.694 | 0.283 | 0.894 | 0.211 | 0.212 | 0.068 | 0.4885 | 0.024 | 0.238 |
