## Supplementary material for "Metagenomic Analysis Reveals A Possible Association Between Respiratory Infection and Periodontitis": Table S10.docx

**Table S10.** **Quality control of dataset by Wang et al. 2016.**

| **Sample** | **Raw reads** | **Clean reads** | **Clean read rate** | **Non-human reads** | **Non-human**  **read rate (%)** |
| --- | --- | --- | --- | --- | --- |
| PDP1 | 12,320,794 | 11,987,568 | 0.972954178 | 66,320 | 0.55% |
| PDP10 | 18,265,188 | 17,776,849 | 0.973263949 | 16,669,388 | 93.77% |
| PDP2 | 11,841,736 | 11,386,534 | 0.961559521 | 10,101,664 | 88.72% |
| PDP3 | 80,73,004 | 77,25,145 | 0.956910835 | 7,064,932 | 91.45% |
| PDP4 | 11,569,812 | 11,106,540 | 0.959958554 | 10,321,458 | 92.93% |
| PDP5 | 5293956 | 5056077 | 0.955065928 | 3795744 | 75.07% |
| PDP6 | 13307752 | 11857595 | 0.891029154 | 8241658 | 69.51% |
| PDP7 | 18519330 | 16852052 | 0.909970933 | 768342 | 4.56% |
| PDP8 | 7600298 | 6458843 | 0.849814441 | 5399996 | 83.61% |
| PDP9 | 8313926 | 7059615 | 0.849131325 | 5510454 | 78.06% |
| PHP1 | 9792960 | 8441360 | 0.861982485 | 1168218 | 13.84% |
| PHP10 | 15275976 | 14378065 | 0.941220712 | 50266 | 0.35% |
| PHP2 | 10574606 | 9103033 | 0.860838976 | 8112546 | 89.12% |
| PHP3 | 8892556 | 7779260 | 0.874805849 | 6908234 | 88.80% |
| PHP4 | 8122434 | 6918064 | 0.851723018 | 5928884 | 85.70% |
| PHP5 | 11931698 | 10349463 | 0.867392302 | 9237360 | 89.25% |
| PHP6 | 15200260 | 13424969 | 0.883206537 | 12144514 | 90.46% |
| PHP7 | 4699594 | 4378227 | 0.931618136 | 2774174 | 63.36% |
| PHP8 | 7605602 | 7403074 | 0.973371207 | 6844780 | 92.46% |
| PHP9 | 3270778 | 3159637 | 0.966020011 | 2539256 | 80.37% |
