## Supplementary figures and images for "Metagenomic Analysis Reveals A Possible Association Between Respiratory Infection and Periodontitis"

### Figure S1.pdf

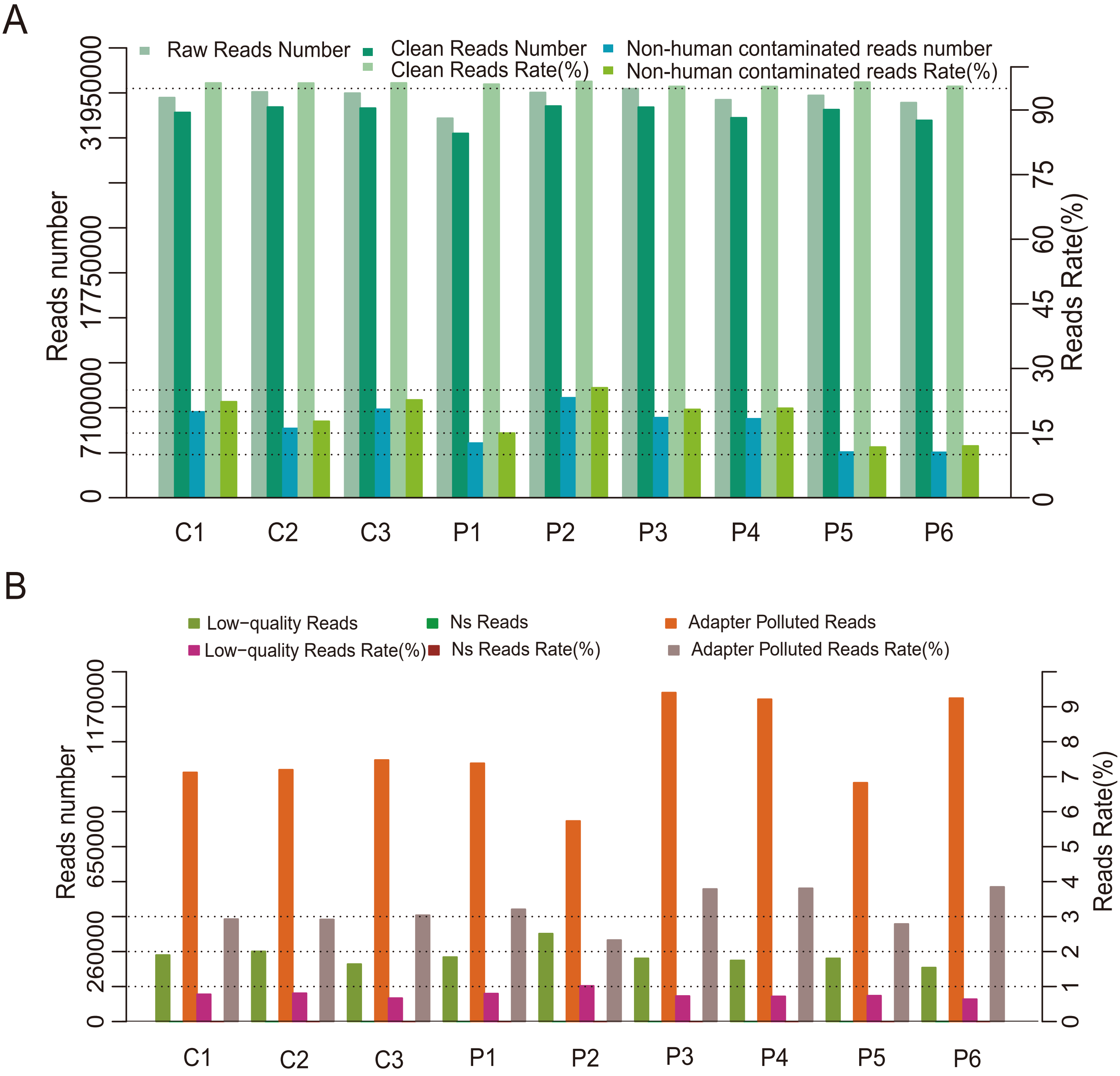

### Figure S2.pdf

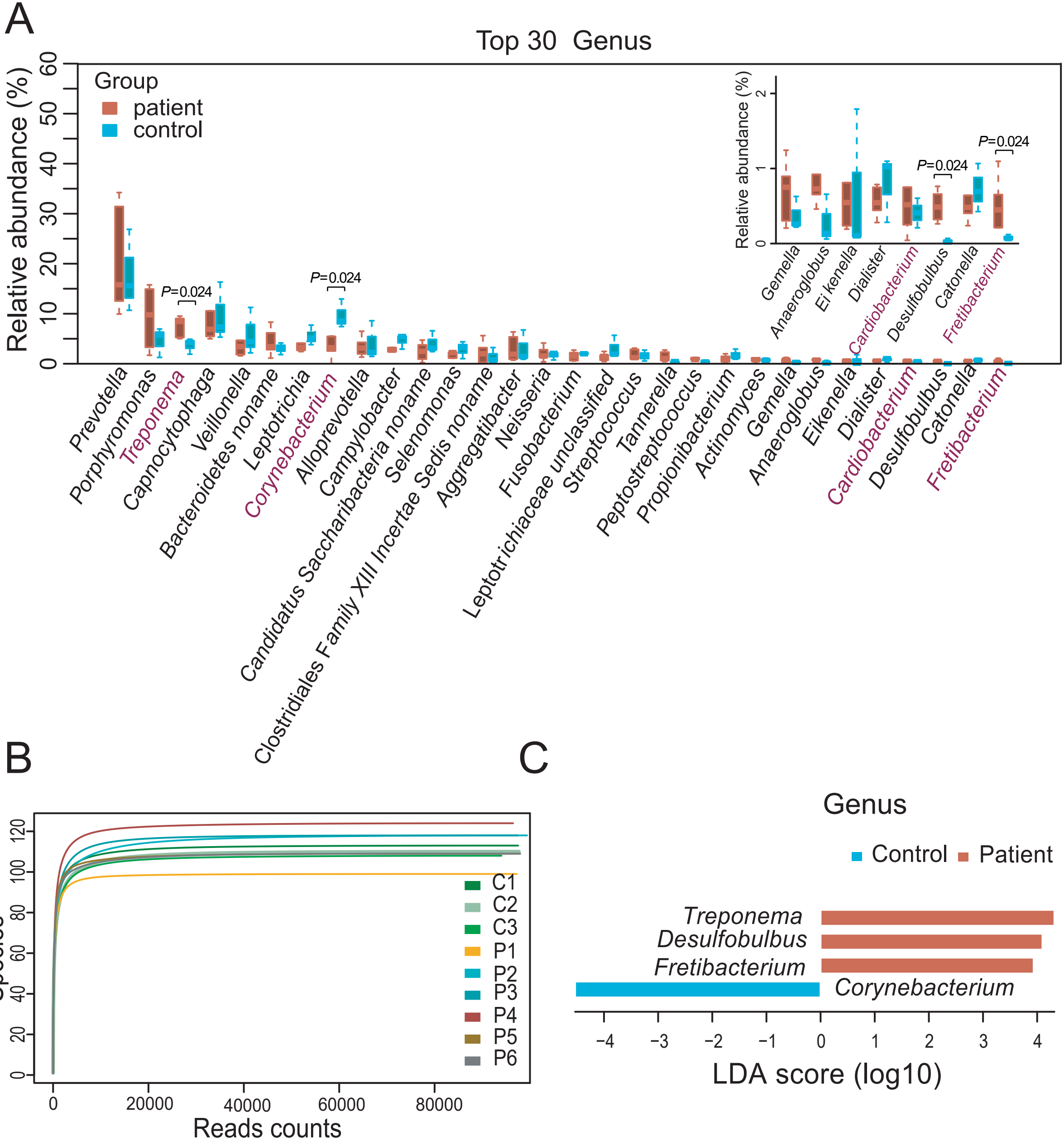

### Figure S3.pdf

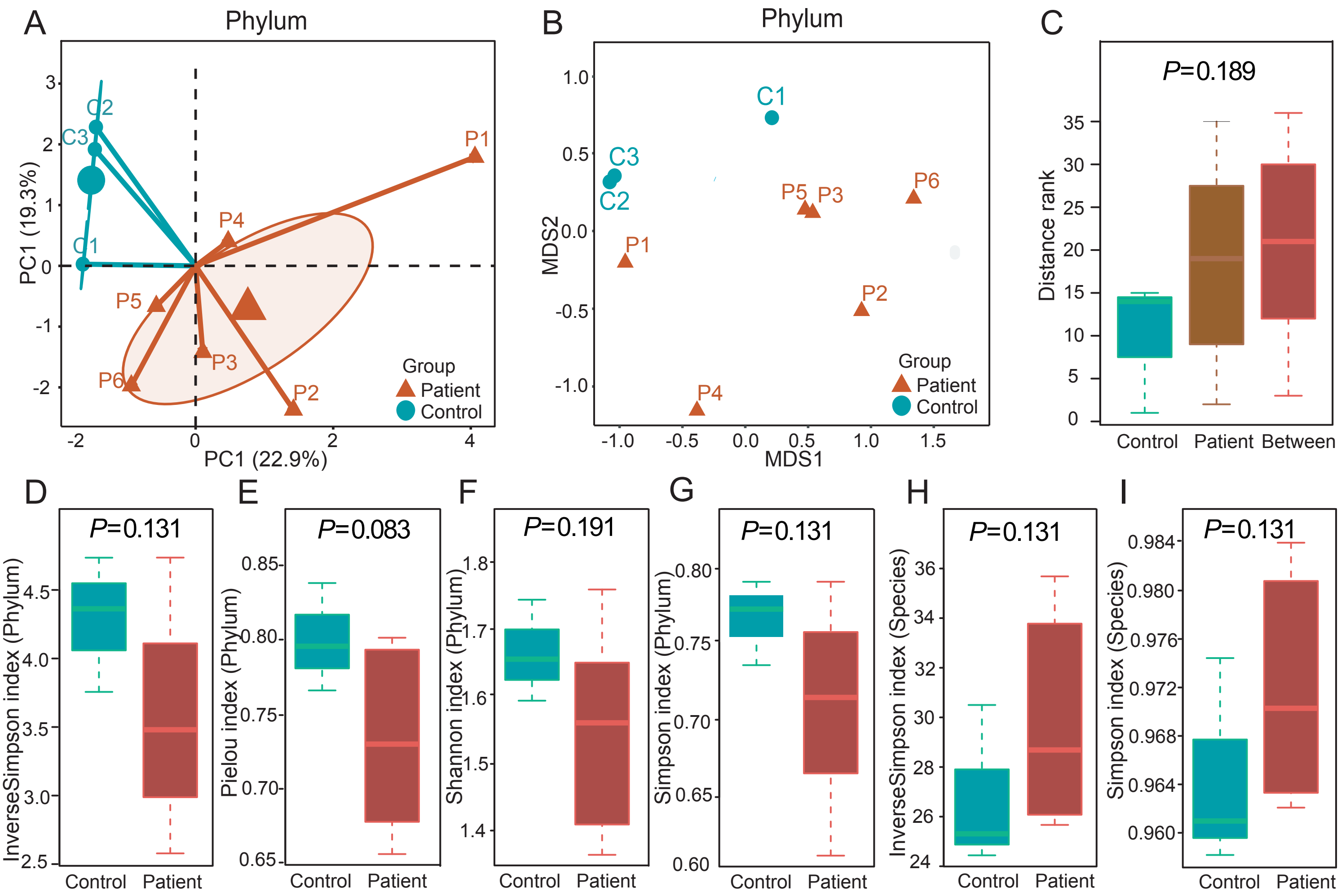

### Figure S4.pdf

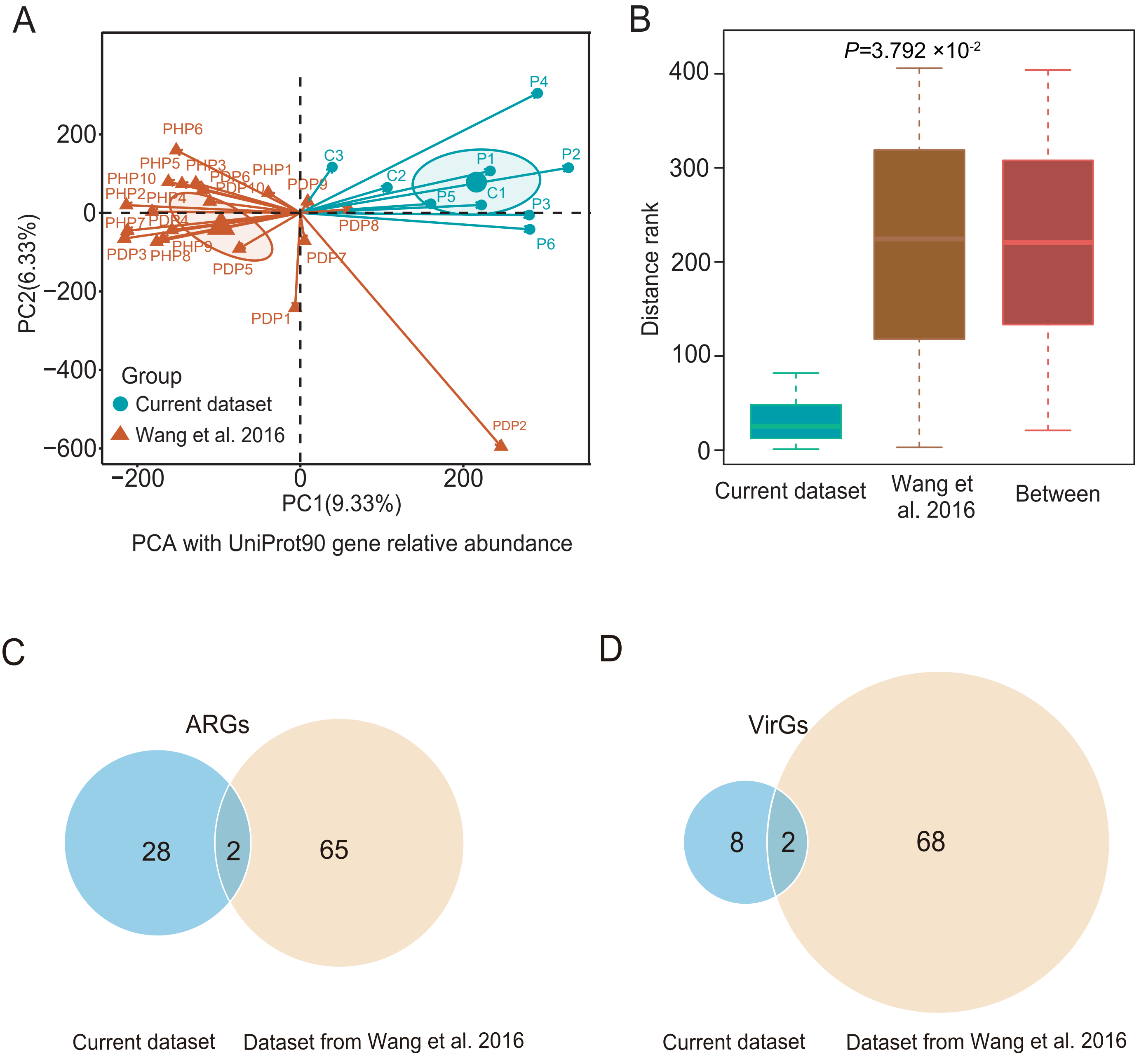

### Figure S5.pdf

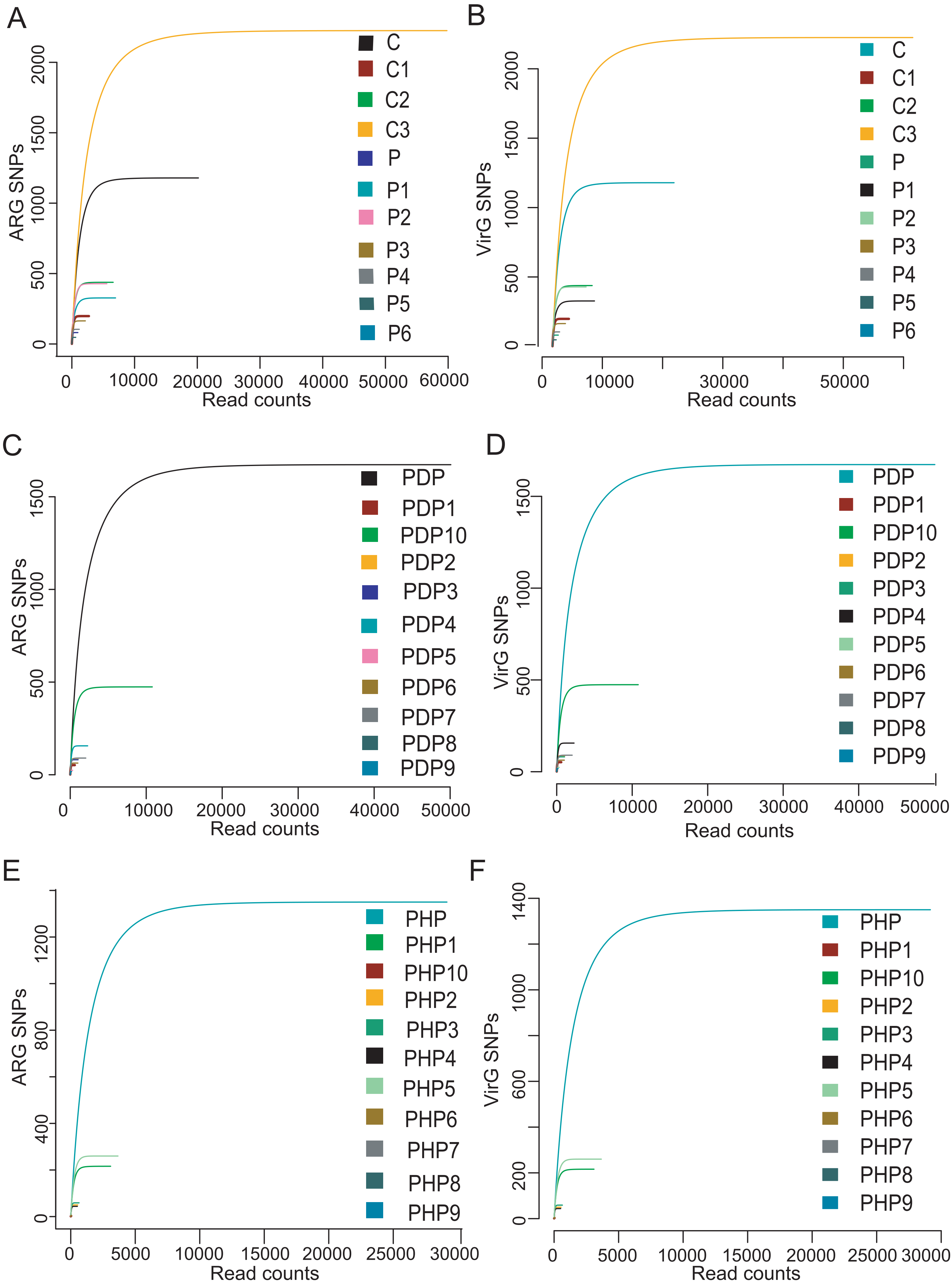

### Figure S6.pdf

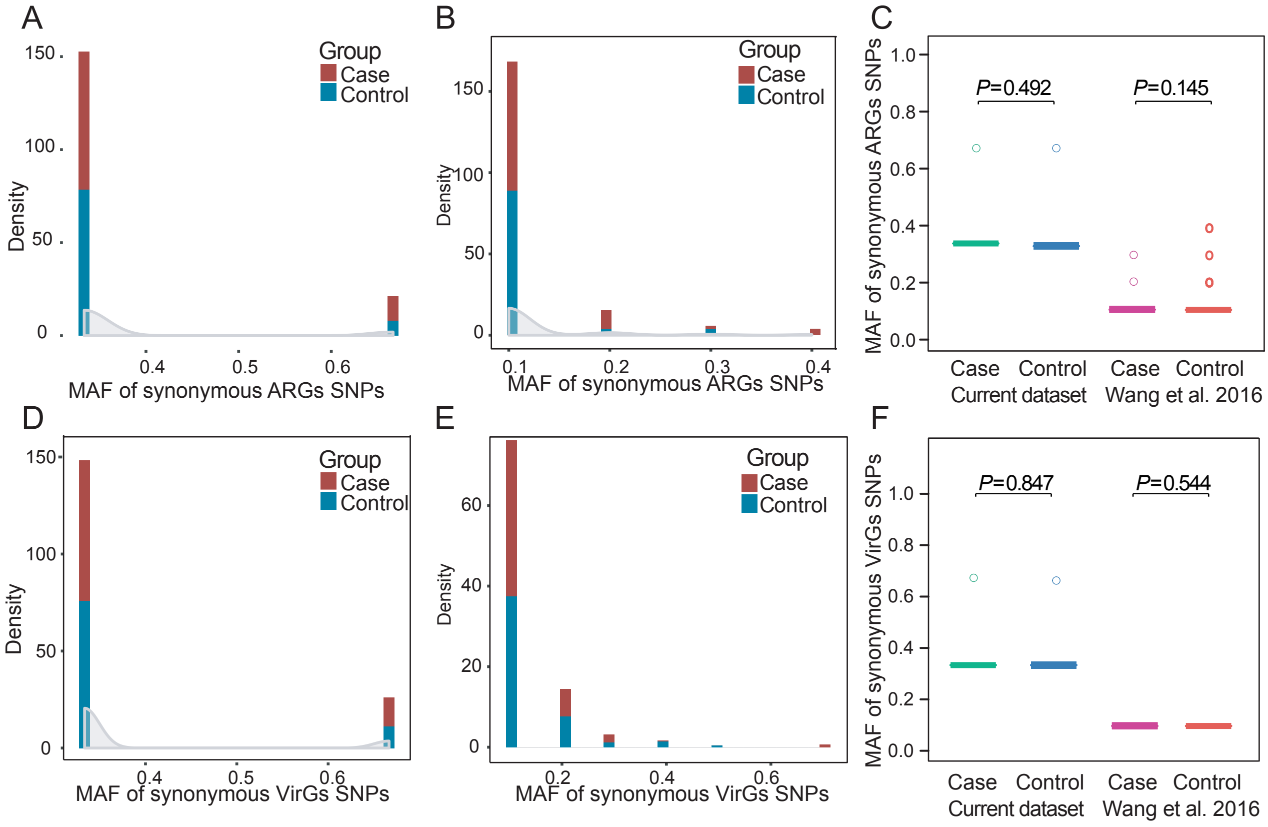
